# Plasma proteomics identifies molecular subtypes in sepsis

**DOI:** 10.1101/2025.05.22.25328197

**Authors:** Thilo Bracht, Kerstin Kappler, Malte Bayer, Franziska Grell, Karin Schork, Lars Palmowski, Björn Koos, Tim Rahmel, Dominik Ziehe, Matthias Unterberg, Lars Bergmann, Katharina Rump, Martina Broecker-Preuss, Ulrich Limper, Dietrich Henzler, Stefan Felix Ehrentraut, Thilo von Groote, Alexander Zarbock, Stephanie Pfaender, Nina Babel, Katrin Marcus-Alic, Martin Eisenacher, Michael Adamzik, Barbara Sitek, Hartmuth Nowak

## Abstract

**Background:** The heterogeneity of sepsis represents a significant challenge to the development of personalized sepsis therapies. Sepsis subtyping has therefore emerged as an important approach to this problem, but its impact on clinical practice was limited due to insufficient molecular insights. Modern proteomics techniques allow the identification of subtypes and provide molecular and mechanistical insights. In this study, we analyzed a prospective multi-center sepsis cohort using plasma proteomics to describe and characterize sepsis plasma proteome subtypes.

**Methods:** Plasma samples were collected from 333 patients at days 1 and 4 of sepsis and analyzed using liquid chromatography coupled to tandem mass spectrometry. Plasma proteome subtypes were identified using K-means clustering and characterized based on clinical routine data, cytokine measurements, and proteomics data. A random forest machine learning classifier was generated to enable future assignment of patients to subtypes.

**Results:** Four subtypes with different sepsis severity were identified. Cluster 0 represented the most severe form of sepsis, with 100 % mortality. Cluster 1, 2 and 3 showed a gradual decrease of the median SOFA score, as reflected by clinical data and cytokine measurements. At the proteome level, the subtypes were characterized by distinct molecular features. We observed an alternating immune response, with cluster 1 showing prominent activation of the adaptive immune system, as indicated by elevated levels immunoglobulin (Ig) levels, which were verified using orthogonal measurements. Cluster 2 was characterized by acute inflammation and the lowest Ig levels. Cluster 3 represented the sepsis proteome baseline of the investigated cohort. We generated an ML classifier and optimized it for the minimum number of proteins that could realistically be implemented into routine diagnostics. The final model, which was based on 10 proteins and Ig quantities, allowed the assignment of patients to clusters 1, 2 and 3 with high confidence.

**Conclusion:** The identified plasma proteome subtypes provide insights into the immune response and disease mechanisms and allow conclusions on appropriate therapeutic measures, enabling predictive enrichment in clinical trials. Thus, they represent a step forward in the development of targeted therapies and personalized medicine for sepsis.

## Introduction

Sepsis is a life-threatening condition caused by a dysregulated immune response to an infection [1]. It is a frequent cause of death in intensive care units (ICU) and represents a major burden in global healthcare with persistently high mortality rates [2]. Up to now, the major improvements in sepsis therapy were based on the effective treatment of the underlying infection and supportive measures [3]. All therapeutic approaches that specifically targeted molecular mechanisms in sepsis failed in large clinical trials [4]. The major reason for this is seen in the fact that sepsis is a complex, multifactorial syndrome and patient subgroups that might benefit from the tested medication were masked by others in heterogeneous clinical cohorts [5]. The concept of personalized medicine is based on the identification and treatment of only those patients that benefit from a therapy. This, however, requires accurate knowledge of the pathophysiological processes underlying the disease and appropriate markers for patient stratification.

In this respect, high throughput technologies can be used to screen patients for biomarkers and elucidate molecular disease mechanisms in an unbiased manner. Mass spectrometry-based plasma proteomics have emerged as a powerful analytical tool, giving access to large patient cohorts and a wide range of blood proteins. Despite these opportunities, studies that analyzed the blood proteome of septic patients remain comparably rare [6–9]. A landmark study published in 2024 was the first with a cohort size that allowed the identification of patient subgroups representing molecular sepsis subtypes [10]. This approach was complementary to others, that mostly used clinical standard parameters or transcriptomic data [11–14]. The generalizability of phenotyping studies, however, remains questionable, especially between different geographic locations [15, 16]. We conclude that beyond the mere identification, the molecular and clinical characterization of subtypes is an absolute necessity to receive transferable insights. In addition, a deep characterization is crucial to elucidate the pathophysiological relevance of the observed subtypes and infer future therapeutic concepts.

In this study, we analyzed proteomics data from a multicenter cohort comprising 333 patients and samples collected at two time points in early sepsis. We applied cluster analysis and identified four sepsis plasma proteome subtypes and their trajectories between both time points. We present a deep characterization of the subtypes and their clinical characteristics. In addition, we developed a supervised machine learning model to allocate new patients to the proteome clusters based on a minimized number of features, demonstrating how proteome subtypes could be implemented in clinical practice in the future.

## Methods

### Patient cohort

Patients with sepsis according to Sepsis-3 definition [1] were enrolled within the multi-center, prospective, observational SepsisDataNet.NRW and CovidDataNet.NRW studies (German Clinical Trial Registry, No. DRKS00018871). The diagnosis of sepsis had to be made within 48 hours before study inclusion, which then was on the first day of treatment in the ICU. Blood samples (EDTA plasma and serum) were collected at day 1 and day 4. Patient-related ICU data and baseline characteristics were derived from the electronic patient’s records. Treatment was provided in accordance with current guidelines and was not influenced by study inclusion.

### Plasma Proteomics

Proteome analyses of plasma samples were conducted as described recently [6]. Briefly, 1 µl of plasma per sample was digested using the SP3 protocol and 545 samples from 333 patients were analyzed distributed over eight batches. Batches C3 and S9 were analyzed on a Vanquish Neo UHPLC coupled to an Orbitrap Exploris 240. Batch S3 was analyzed using an Ultimate 3000 RSLCnano HPLC coupled to an Orbitrap Fusion Lumos mass spectrometer. All other batches were measured on an Ultimate 3000 RSLCnano HPLC coupled to an Orbitrap Exploris 240 (all Thermo Scientific, Bremen, Germany). The samples were measured in data-independent acquisition mode and data were processed using DIA-NN (v.1.8.1) searching the UniProt/SwissProt database restricted to *Homo sapiens* (v.2022_05). All batches were processed separately and subsequently normalized and evaluated as described before [17] (Supplementary Figure 1). Functional annotation and enrichment analyses were performed using the STRING web interface (string-db.org, v.12.0).

### Proteome Clustering

Clustering was performed using day 1 protein intensities. Only proteins measured at both timepoints (days 1 and 4) with less than 30% missing values at each timepoint were included in the analysis. For clustering, missing values were imputed with the median. First, day 1 protein intensities were standardized using z-transformation for each protein. The standardization parameters (mean and standard deviation) were then applied to day 4 data to ensure consistent transformation across the timepoints. Subsequently, a Principal Component Analysis (PCA) was calculated for day 1 to reduce dimensionality. The resulting principal components were used to project day 4 data into the same reduced feature space. Sepsis plasma proteome subtypes were identified using the k-means algorithm on the PCA results of the first 35 components (>70% of variance). The optimal number of clusters was determined using silhouette curves and initial biological interpretation of the results, resulting in four distinct clusters (Supplementary Figure 2). To assign day 4 data to the clusters, the Euclidean distance to each cluster centre was calculated, and each data point was assigned to the cluster with the nearest cluster centre. All calculations were performed using Python (v.3.10.14) and the packages pandas (v.2.1.1) and scikit-learn (v.1.3.1).

### Cytokine measurements

The LegendPlex Human Inflammation Panel 1 (Biolegend, San Diego) was used according to the manufacturer’s instructions as described previously [18]. Briefly, serum samples were mixed and incubated with the LegendPlex beads, subsequently washed, and incubated with detection antibodies. After a final washing step, the measurements were carried out using a flow cytometer (Canto II, BD Biosciences, CA). Cytokine concentrations were interpolated from calibration curves that were generated and measured together with the analyzed samples. Calculated concentrations below the lower limit of quantification (LOD) were considered to be zero, concentrations higher than the upper LOD were replaced by the respective upper LOD.

### Immunoglobulin quantification

Serum aliquots were thawed and centrifuged at 10000xg for 5 seconds to pellet precipitates. The supernatant was used to determine the concentration of IgG, IgA and IgM. The measurement was performed turbidimetrically on the Cobas c501 system (Roche Diagnostics, Mannheim, Germany) using Roche reagents (Tina-quant IgA, IgG and IgM, second gen.), according to the manufacturer’s instructions.

### Statistics

Clinical variables with multiple observations per day were aggregated by the median, minimum or maximum using the most clinical informative value (e.g. minimum value for quick value and maximum value for creatinine). The 25th percentile, median, and 75th percentile were calculated for each variable and subtype. For categorical variables, p-values were calculated using the chi-squared or fisher test and adjusted using Bonferroni correction. Kruskal-Wallis followed by Dunn’s post-hoc test for pairwise comparisons was applied for continuous variables. Differences in protein intensities between the four proteome subtypes were tested for statistical significance using ANOVA followed by Tukey’s post-hoc test. ANOVA p-values were corrected according to Benjamini-Hochberg. Relative changes between the clusters were calculated as ratios of mean protein intensities. The significance threshold was set as an ANOVA p_FDR_ value ≤ 0.05, a posthoc-test p value ≤ 0.05 and a ratio of means (RoM) ≥ 1.5 or ≤ 0.67.

### Machine Learning

The proteomics data set was filtered for completeness as described above. In favour of quantities determined by clinical routine diagnostics, all entries representing immunoglobulins were removed and replaced by IgG, IgA, and IgM concentrations that were determined orthogonally. Proteins represented by routine measurements such as C-reactive protein (CRP), Fibrinogen gamma chain (FGG), Fibrinogen alpha chain (FGA), Fibrinogen beta chain (FGB), Hemoglobin subunit alpha (HBA2,) Hemoglobin subunit beta (HBB), and Hemoglobin subunit delta (HBD) were excluded as well (Supplementary Table 1). For the clinical dataset, routine ICU data with less than 30% missing values was considered. Variables with multiple observations per day were aggregated as described above and missing values were completed using data from the subsequent two days. Before model development, highly correlated features were removed by calculating the Pearson correlation between features and excluding one of the two features with an absolute correlation greater than 0.7 (Supplementary Table 2). For clinical data, only variables with a Kruskal–Wallis test *p*-value < 0.05 were used (Supplementary Tables 3 and 4). Cluster 0 was not considered for ML due to its low patient count, which was insufficient for robust modelling.

To identify the most relevant features, a multiclass classifier based on a Random Forest model was employed (Supplementary Figure 3). The data were split into training and test sets (80 % training, 20 % test), and a 100 times Monte Carlo cross-validation (MCCV) was performed to evaluate the model metrics for different feature sets. Feature selection was conducted in a nested iterative manner: medical and proteomic features were added separately in two distinct loops. In the outer loop, medical features were incrementally included based on their importance ranking, while in the inner loop, proteomic features were iteratively added for each medical feature subset. This approach enabled independent evaluation of features from each domain. The order of features for iterative training was determined based on an additional 100 times MCCV (80 % training, 20 % validation). In this process, the median ranks of feature importance were computed using SHAP values (Shapley Additive Explanations) for medical and proteomics data. Missing values were imputed using the median in each MCCV iteration. The point at which the recall growth rate markedly declined was identified using the *KneeLocator* function of the keed package in Python. Based on this threshold, the most frequently selected proteins across MCCV iterations and IgA, IgG and IgM were used to train a final Random Forest model (Figure 3b), for which SHAP values were calculated to assess the contribution of each feature to the model’s predictions. The applied hyperparameters can be found in Supplementary Table 5. Calculations were done using Python (v.3.10.12) and the packages pandas (v.2.2.2), numpy (v.1.26.4), keed (v.0.8.5), shap (v0.46.0) and scikit-learn (v.1.5.1).

## Results

### Cohort characteristics

The cohort comprised n = 333 patients that were enrolled in five university hospitals in Germany. The median age was 64 years (IQR 55-75) with 117 females (37.9 %) and 216 males (62.1 %). 74 patients (24.3 %) showed a SARS-CoV-2 infection and the median SOFA score was 9 (IQR 5-12). The 30-day mortality was 39.6 % (n = 132 patients). For all patients, plasma proteome analyses were done for day 1 while measurements for day 4 plasma samples were available for 213 patients.

### Identification, trajectories and clinical representation of the sepsis plasma proteome subtypes

The k-means algorithm identified four sepsis plasma proteome subtypes on data from day 1. The four clusters showed a decreasing severity of sepsis as represented by the SOFA score with the highest scores found in cluster 0 and the lowest in cluster 3 (Figure 1a, Table 1). Consequently, the highest 30-day mortality was also observed in cluster 0 (Figures 1b and c), while no significant differences in 30-day mortality were found between the other three clusters. The median survival time in cluster 0 was 2 days at day 1 (IQR 0 - 3) and 4 days at day 4 (IQR 3 – 4.5). No significant differences were found for age, sex and the focus of infection (Supplementary Table 3). Proteome data corresponding to day 4 were used to assign the respective patients to the same clusters. The results illustrate a relative stability of the clusters with the majority of patients staying in one cluster. Some patients, however, also migrated from one cluster to another. Migration to cluster 0 showed a striking relationship with the outcome, with all patients passing through cluster 0 eventually dying (Figure 1b). Patients in cluster 0 showed significantly shorter length of the ICU and hospital stay (Supplementary Figure 4). Clinically, cluster 0 was characterized by acute liver failure (AST, ALT, LDH), lactate acidosis and accompanying high-grade acute kidney injury (AKI). Furthermore, aPPT, INR, platelet count and fibrinogen concentration indicated disseminated intravascular coagulation (DIC). The decrease of CRP due to excessive consumption was less pronounced on day 4 compared to day 1 (Figure 1d, Supplementary Table 3, Supplementary Figure 5). These characteristics underlined the moribund character of patients in cluster 0.

**Figure 1.**
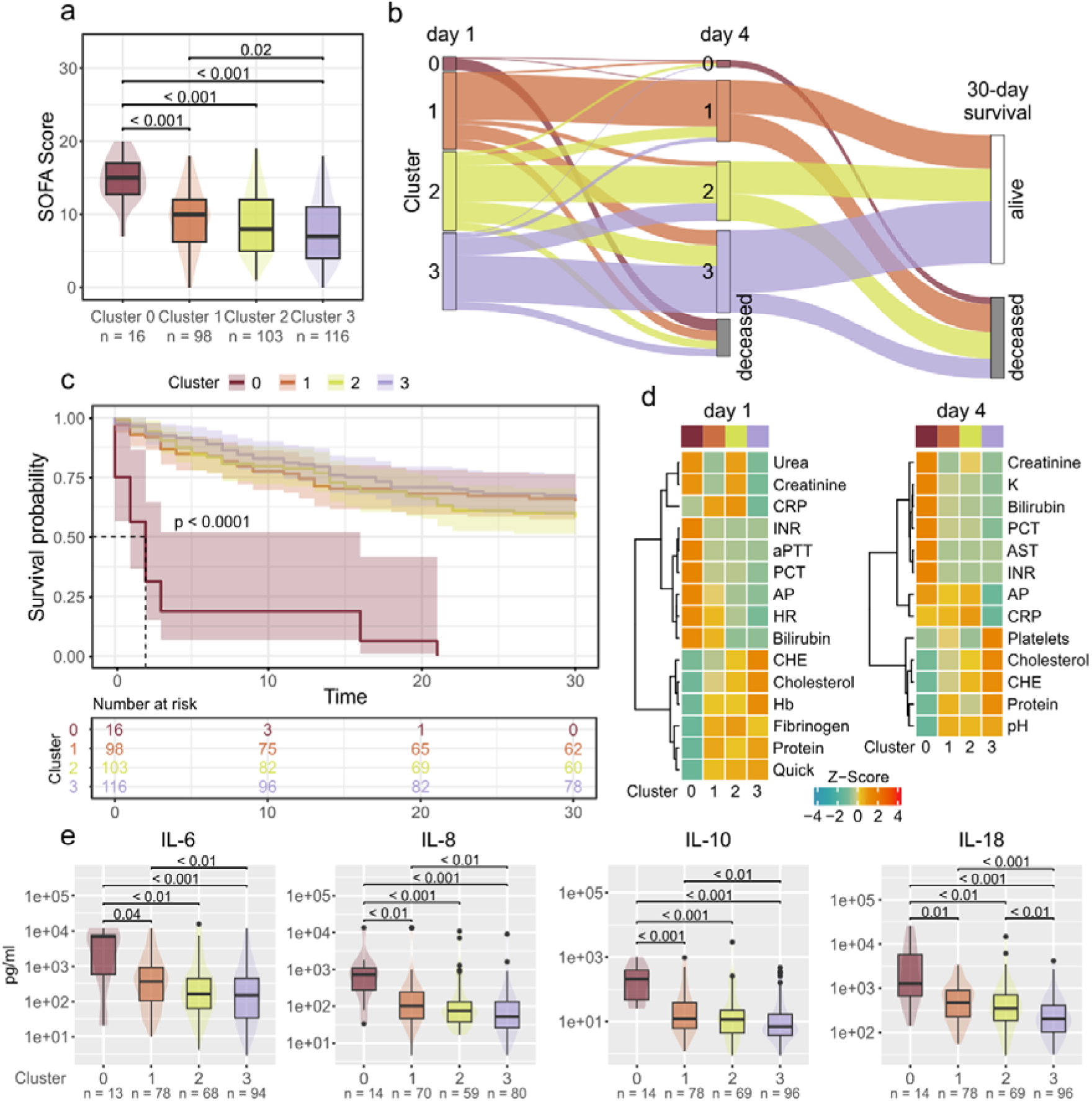
Clinical characteristics of sepsis plasma proteome subtypes. **a** Distribution of SOFA scores across the four clusters shown as boxplots. Boxes represent 25th and 75th percentiles, whiskers extend to the most extreme data points, median shown as a horizontal line, p-values from Dunn’s post-hoc test. **b** Sankey diagram showing the assignment of patients to proteome subtypes at day 1 and day 4, respectively. The endpoint is the 30-day survival. **c** Kaplan-Meier analysis according to 30-day survival for the four subtypes. **d** Heatmaps illustrating clinical routine data at day 1 and day 4. Only parameters which were significant between the clusters 1, 2 or 3 are shown (Bonferroni corrected Kruskal-Wallis followed by Dunn’s test). Data was aggregated by the mean, z-transformed, and clustered using Euclidean distance with Ward’s linkage method. **e** Boxplots illustrating cytokine measurements at day 1. Boxes represent 25th and 75th percentiles, whiskers extend to the most extreme data points, median shown as a horizontal line, outliers shown as individual data points, p-values from Dunn’s post-hoc test.

**Table 1:**
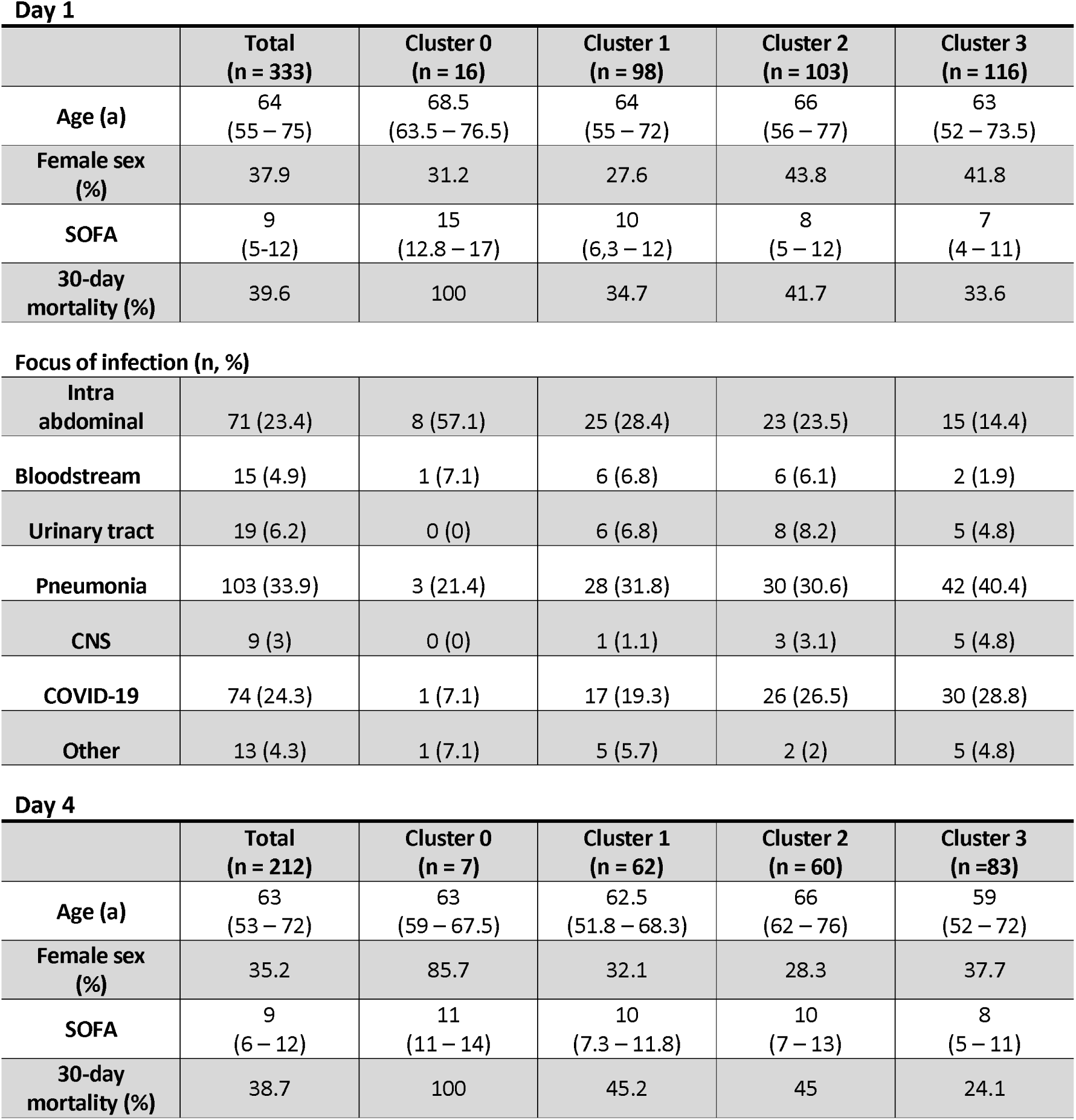
Baseline characteristics of the studied patient cohort, separated for the identified sepsis plasma proteome subtypes.

**Day 1**
|  | **Total**<br>**(n = 333)** | **Cluster 0**<br>**(n = 16)** | **Cluster 1**<br>**(n = 98)** | **Cluster 2**<br>**(n = 103)** | **Cluster 3**<br>**(n = 116)** |
| --- | --- | --- | --- | --- | --- |
| **Age (a)** | 64<br>(55 – 75) | 68.5<br>(63.5 – 76.5) | 64<br>(55 – 72) | 66<br>(56 – 77) | 63<br>(52 – 73.5) |
| **Female sex (%)** | 37.9 | 31.2 | 27.6 | 43.8 | 41.8 |
| **SOFA** | 9<br>(5-12) | 15<br>(12.8 – 17) | 10<br>(6,3 – 12) | 8<br>(5 – 12) | 7<br>(4 – 11) |
| **30-day mortality (%)** | 39.6 | 100 | 34.7 | 41.7 | 33.6 |
| **Focus of infection (n, %)** |  |  |  |  |  |
| **Intra abdominal** | 71 (23.4) | 8 (57.1) | 25 (28.4) | 23 (23.5) | 15 (14.4) |
| **Bloodstream** | 15 (4.9) | 1 (7.1) | 6 (6.8) | 6 (6.1) | 2 (1.9) |
| **Urinary tract** | 19 (6.2) | 0 (0) | 6 (6.8) | 8 (8.2) | 5 (4.8) |
| **Pneumonia** | 103 (33.9) | 3 (21.4) | 28 (31.8) | 30 (30.6) | 42 (40.4) |
| **CNS** | 9 (3) | 0 (0) | 1 (1.1) | 3 (3.1) | 5 (4.8) |
| **COVID-19** | 74 (24.3) | 1 (7.1) | 17 (19.3) | 26 (26.5) | 30 (28.8) |
| **Other** | 13 (4.3) | 1 (7.1) | 5 (5.7) | 2 (2) | 5 (4.8) |

**Day 4**
|  | **Total**<br>**(n = 212)** | **Cluster 0**<br>**(n = 7)** | **Cluster 1**<br>**(n = 62)** | **Cluster 2**<br>**(n = 60)** | **Cluster 3**<br>**(n = 83)** |
| --- | --- | --- | --- | --- | --- |
| **Age (a)** | 63<br>(53 – 72) | 63<br>(59 – 67.5) | 62.5<br>(51.8 – 68.3) | 66<br>(62 – 76) | 59<br>(52 – 72) |
| **Female sex (%)** | 35.2 | 85.7 | 32.1 | 28.3 | 37.7 |
| **SOFA** | 9<br>(6 – 12) | 11<br>(11 – 14) | 10<br>(7.3 – 11.8) | 10<br>(7 – 13) | 8<br>(5 – 11) |
| **30-day mortality (%)** | 38.7 | 100 | 45.2 | 45 | 24.1 |

Cluster 1 showed a higher heart rate and elevated liver function markers AP, bilirubin, as well as lower CHE and cholesterol at day 1. On day 4, the heart rate was not significantly different between the four clusters, which was likely an effect of stabilization by the applied therapy. On day 1, cluster 2 was characterized by a moderate AKI with higher creatinine and urea levels and had the highest CRP levels of all four subtypes. Total protein was lower compared to clusters 1 and 3, an observation that was more pronounced on day 4, when cluster 2 also showed elevated AP levels and a decreased platelet count. Cluster 3 showed the overall mildest sepsis phenotypes and clinical values that were least alarming. Thus, it may be considered a baseline in our data set, however, the 30-day survival was not significantly different from clusters 1 and 2.

### Immune status

At day 1, we found significant differences in the levels of IL-6, IL-8, IL10, IL-18 and CCL-2 (Supplementary Table 3). The distribution of cytokine concentrations resembled the distribution of the SOFA score, with the highest levels observed in cluster 0 and lowest in cluster 3 (Figure 1e). At day 4, we additionally found IL-17a to be significantly differential between clusters 1 and 3. The counts of lymphocytes and white blood cells were not significantly differential between the subtypes.

### Characteristics of the Sepsis Plasma Proteome Subtypes

Proteome analysis revealed the greatest differences between the subtypes in comparison to cluster 0. Overall, we found 271 significantly differential proteins between all four clusters at day 1 and 206 proteins at day 4 (Figure 2a, Supplementary Figure 6, Supplementary Table 6). Of these, 247 and 181 proteins were differential in comparison to cluster 0 at day 1 and day 4, respectively. This illustrates the unique plasma composition of cluster 0 which was associated with the most severe sepsis and highest mortality. The hierarchical cluster analysis of all significant proteins revealed five characteristic abundance patterns in relation to the identified subtypes on day 1. On day 4, four patterns could be discriminated, with regulation pattern α missing (Supplementary Figures 6 and 7, Supplementary Table 7).

**Figure 2.**
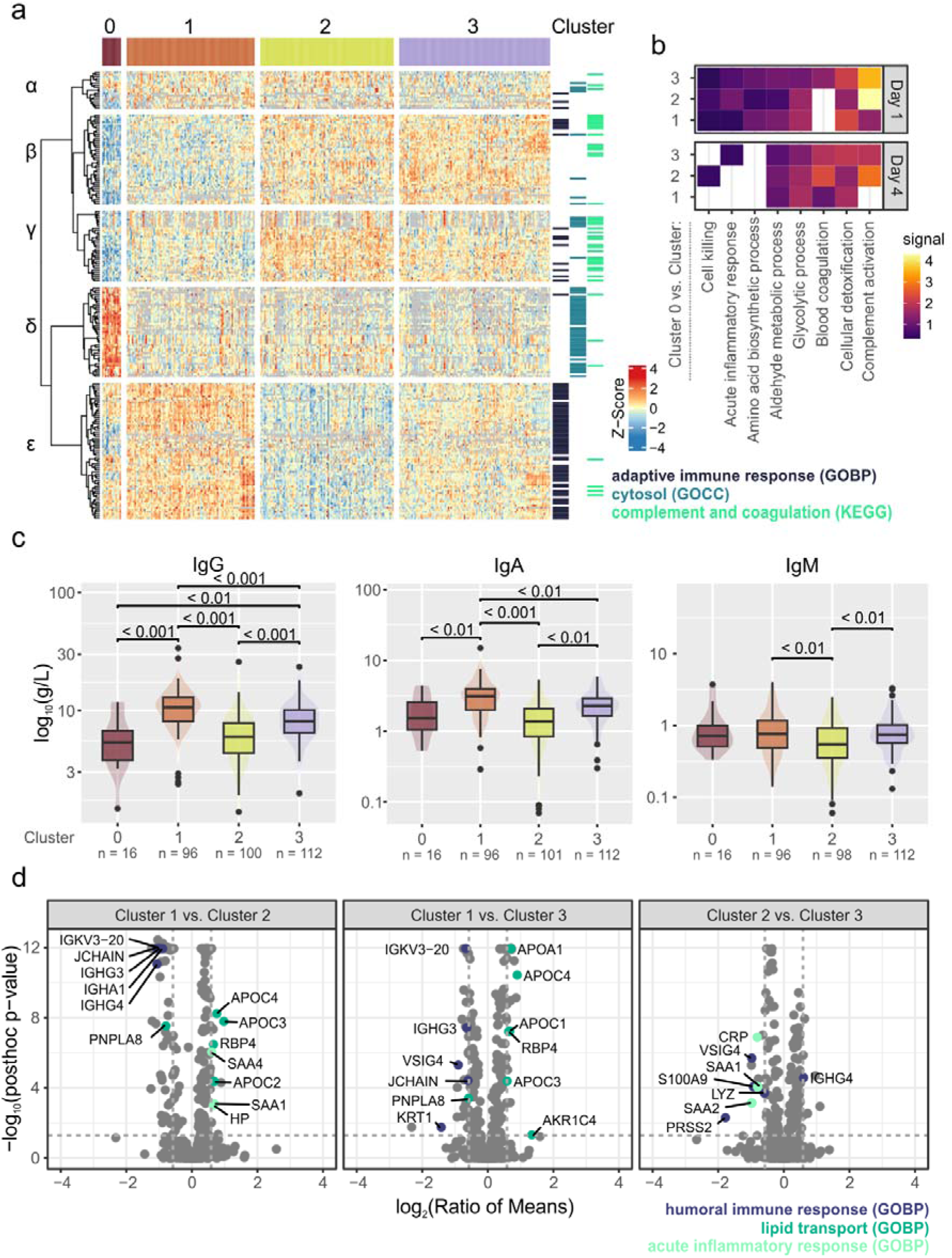
Proteome characteristics of sepsis plasma proteome subtypes. **a** Heatmap showing significantly differentially abundant proteins at day 1 (ANOVA pFDR value ≤ 0.05, post-hoc test p value ≤ 0.05, ratio of means ≥ 1.5 or ≤ 0.67). Protein intensities were z-transformed and clustered using Pearson clustering with Ward’s linkage method. The first five branches in the dendrogram were divided and labeled with Greek letters to discriminate abundance patterns. Protein annotation with selected Gene Ontology or KEGG categories is indicated on the right. **b** Functional enrichment of significantly differential proteins in comparison to cluster 0. Eight selected categories shown for day 1 and day 4 and the three pairwise comparisons with cluster 0. Enrichment analysis was done with string-db.org (v.12) using GO biological processes (GOBP). **c** Immunoglobulin measurements as boxplot representation. Boxes represent 25th and 75th percentiles, whiskers extend to the most extreme data points, median shown as a horizontal line, outliers shown as individual data points, p-values calculated by Dunn’s post-hoc test. **d** Volcano plots illustrating the pairwise comparisons between clusters 1, 2 and 3 for day 1. Proteins annotated with significantly enriched GOBP terms were highlighted and labeled with gene names. Dashed lines indicate the applied significance threshold.

Typical of cluster 0, we found markers of cell death and tissue damage to be elevated, characterized by their common cytosolic location (Figure 2a, abundance pattern δ). These consisted mainly of liver proteins associated with metabolic processes such as glucose, aldehyde and amino acid metabolism as well a cellular detoxification (Figure 2b). The enriched biological processes might indicate an accumulation of the corresponding proteins before cell death, pointing towards enhanced elimination of reactive oxygen species and a metabolic shift towards anaerobic glycolysis. On the other hand, we found proteins related to blood coagulation but also complement activation and acute phase response to be depleted in cluster 0, showing the lowest levels across all subtypes (abundance patterns β and γ).

Decreasing levels of proteins associated with complement and coagulation as well as acute phase response were also apparent in cluster 1, highlighting the relevance of the associated processes in sepsis. Similarly, apolipoproteins, especially related to remodeling of high-density lipoprotein particles, were lower abundant in cluster 1 compared to clusters 2 and 3 (Figure 2d, Supplementary Figure 6 b and c), showing even more pronounced depletion in cluster 0 (abundance pattern β). This finding was in accordance with clinical values, which indicated reduced liver function in cluster 1 and acute liver failure in cluster 0 (Figure 1d). Another striking finding was the elevation of immunoglobulins (Ig) in cluster 1, in particular because this observation could not be anticipated based on clinical routine data (Figure 2a, abundance pattern β). High levels of immunoglobulins indicated a strong adaptive immune response in this subtype, corresponding to high SOFA scores and cytokine levels. The lowest immunoglobulin levels, in contrast, were found in cluster 2 which also corresponds to characteristically low levels in total protein. As no clinical routine parameter could be used to specifically substantiate these observations, total levels of IgG, IgA and IgM were determined post-hoc. These independent measurements verified the complementary levels of IgG and IgA, and showed the lowest levels of IgM in cluster 2 (Figure 2c). In cluster 2 however, the highest levels of proteins related to acute inflammatory response were observed, including Serum amyloid A1 and A2 (SAA1, SAA2) as well as Haptoglobin (HP), and on day 4 also CRP (Figure 2a, abundance pattern γ, Figure 2d). Within the clusters 1, 2 and 3, the highest levels of the alarmins S100 A8 and A9 were also found in cluster 2 (Supplementary Figure 6).

On day 1, an additional regulation pattern of proteins was observed, showing their lowest levels in cluster 3 (Figure 2a, α pattern). This set of proteins contained proteins related to leukocyte transmigration (ICAM1, VCAM1), the kidney function marker Cystatin C (CST3) and immunoregulatory proteins such as VSIG4. A similar regulation pattern was not observed on day 4. The lowest abundance in cluster 3 as well as the increasing levels corresponding with increasing disease severity underlined the baseline character of cluster 3.

### Prediction of Sepsis Plasma Proteome Subtypes by Supervised Machine Learning

A random forest machine learning (ML) classifier was generated to showcase patient stratification according to sepsis plasma proteome subtypes. First, the number of features beyond which only a marginal improvement in recall was achieved was identified by iteratively adding features to the model and evaluating recall (Figure 3a). Over 100 iterations of Monte Carlo cross-validation (MCCV), the most frequently selected proteins in combinations with ten features were encoded by the following genes: C9, ITIH3, ALB, GPLD1, C2, SERPINF2, C18orf63, SERPINC1, SERPINA4, and PON1 (Table 2, Supplementary Figure 8). At this level, the inclusion of clinical features resulted in only minimal performance gains. We then used the identified features to train a new model and evaluate its metrics by MCCV. As Igs provided particularly relevant information for differentiation of clusters 1 and 2, they were also incorporated into the final model. Here, a discrepancy greater than 0.1 was observed between the training and test sets for all evaluated metrics, indicating overfitting, likely caused by an insufficient number of data points. Despite overfitting, an acceptable model performance was still achieved for test data across all three clusters with an AUROC of 0.932 ± 0.022 as well as a precision of 0.703 ± 0.066 and a recall of 0.802 ± 0.044 (Table 3). We assessed the contribution of individual features to the model predictions and found C9, C2 and SERPINF2 (Figure 3b) with the overall highest predictive values. In Cluster 1, the most important features were ALB, GPLD1, SERPINC1 and SERPINA4 (Figure 3c), which were associated with lower values, while C18orf63 levels were elevated (Supplementary Figure 9). In Cluster 2 the most important features C9, C2, SERPINF2 and ITIH3 showed increased levels. In Cluster 3, ITIH3 and SERPINA4 showed the highest importance, with lower levels of ITIH3 and higher levels of SERPINA4 being associated with cluster-specific predictions.

**Table 2.** Gene and proteins names of the selected biomarker panel.

| Gene | UniProt ID | Protein Name |
| --- | --- | --- |
| ALB | P02768 | Albumin |
| C18orf63 | Q68DL7 | Uncharacterized protein C18orf63 |
| C2 | P06681 | Complement C2 |
| C9 | P02748 | Complement component C9 |
| GPLD1 | P80108 | Phosphatidylinositol-glycan-specific phospholipase D |
| ITIH3 | Q06033 | Inter-alpha-trypsin inhibitor heavy chain H3 |
| PON1 | P27169 | Serum paraoxonase/arylesterase 1 |
| SERPINA4 | P29622 | Kallistatin |
| SERPINC1 | P01008 | Antithrombin-III |
| SERPINF2 | P08697 | Alpha-2-antiplasmin |

**Table 3.**
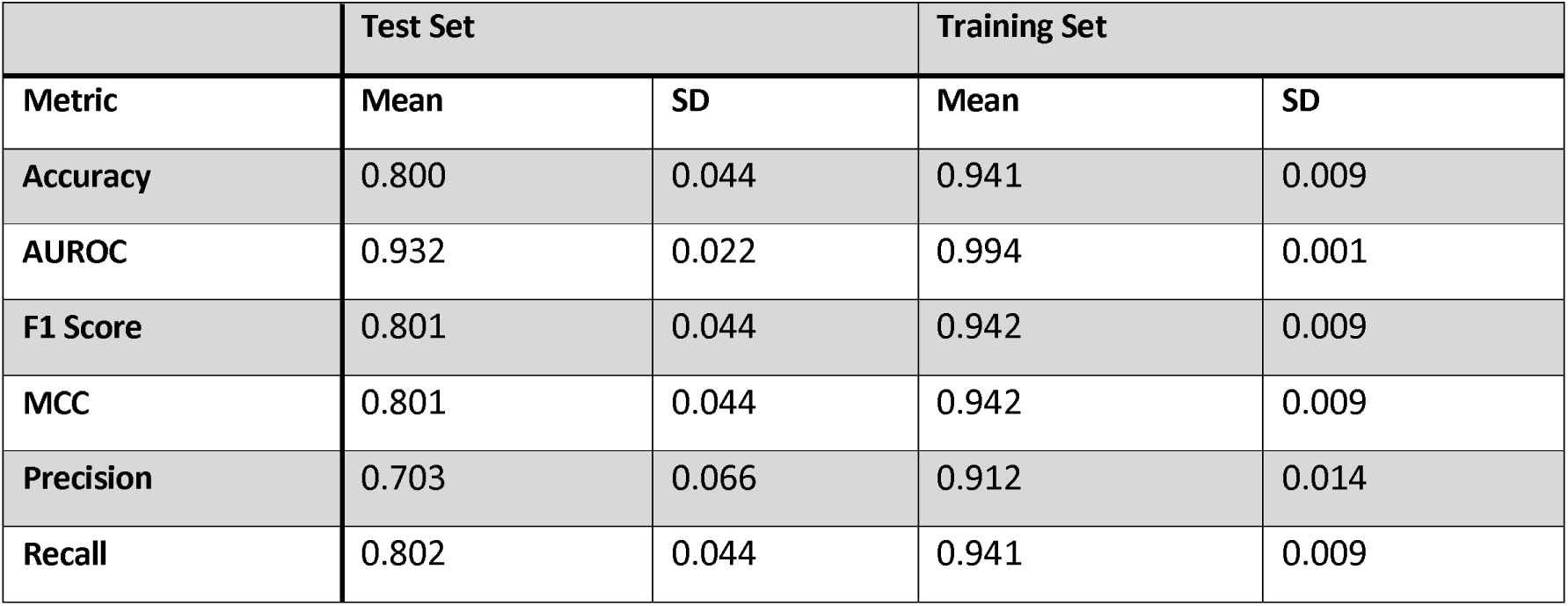
Performance metric of the random forest ML classifier. The classifier was based on 10 proteins and additional Ig quantities and evaluated using 100 times Monte Carlo cross validation.

|  | Test Set |  | Training Set |  |
| --- | --- | --- | --- | --- |
| Metric | Mean | SD | Mean | SD |
| Accuracy | 0.800 | 0.044 | 0.941 | 0.009 |
| AUROC | 0.932 | 0.022 | 0.994 | 0.001 |
| F1 Score | 0.801 | 0.044 | 0.942 | 0.009 |
| MCC | 0.801 | 0.044 | 0.942 | 0.009 |
| Precision | 0.703 | 0.066 | 0.912 | 0.014 |
| Recall | 0.802 | 0.044 | 0.941 | 0.009 |

**Figure 3.**
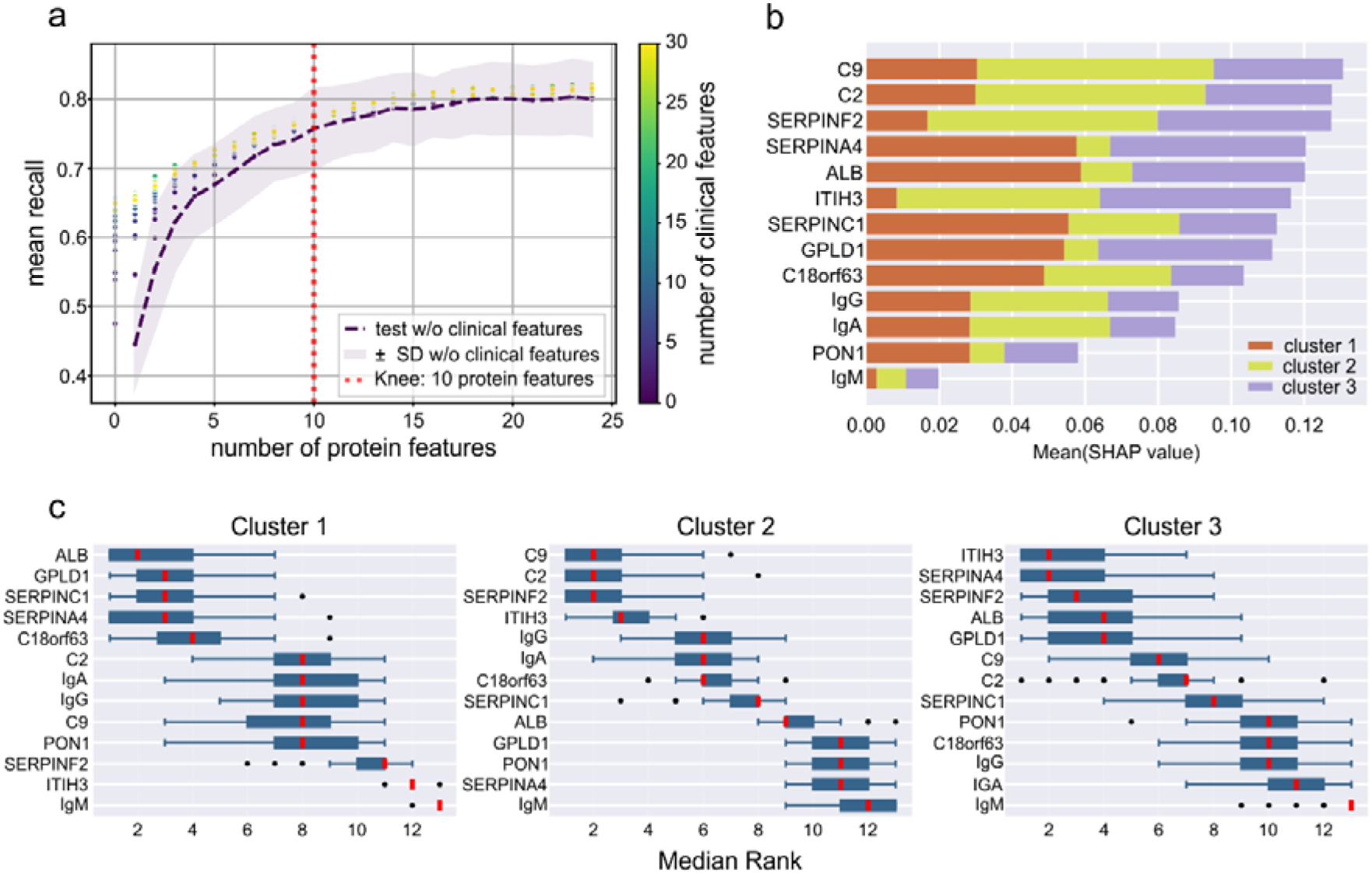
Feature selection and machine learning model performance: **a** Scatter plots representing the mean recall as a function of the number of medical features included. The purple dashed line shows the mean recall when using only protein features. The transparent area curve represents the standard deviation. The red dotted line indicates the computed knee point, where the slope of the curve decreased significantly. With only few proteins available, medical features had a stronger impact on model performance, with increasing number of proteins this effect diminished. **b** Bar plot showing the mean SHAP values for the features used in model training. The colours indicate the contribution of each feature to the respective clusters. **c** Box plots of feature ranks for each cluster, based on the relevance of the features within the model across all MCCV iterations. Boxes represent 25th and 75th percentiles, whiskers extend to the most extreme data points, median shown as a red line, outliers shown as individual data points

## Discussion

The identified plasma proteome subtypes were characterized by different sepsis severity as reflected by the SOFA score and cytokine quantities. Cluster 0 represented the most severe patients with shortest survival and 100 % mortality. This was accompanied by clinical routine data which showed overall extreme values, pointing towards liver failure, acute kidney injury and DIC. Based on these features, cluster 0 seemed to represent rather a terminal stage in sepsis progression than a molecular subtype in the sense of a disease endotype. A significantly shorter time of ICU and hospital stay for patients in cluster 0, however, indicated a rapid worsening that was characteristic to this subtype (Supplementary Figure 4). We conclude that a finer temporal resolution might clarify which patients migrate through cluster 0 when dying, and if it represents an endotype, a time point in sepsis or a mixture of both.

For the other three clusters, we observed striking evidence of different immune responses, with clusters 1 and 2 showing opposite phenotypes. Cluster 1 was characterized by a comprehensive immune reaction, including the adaptive immune system as indicated by high Ig levels. Decreasing complement and coagulation factors as well as acute phase proteins also suggested activation of the innate immune system and consumption of the corresponding proteins. Cluster 2, in contrast, showed the lowest Ig levels but highest levels of acute phase proteins. In addition, elevated intensities of S100A8/A9 proteins hinted towards a pronounced neutrophil degranulation. The immunomodulatory protein VSIG4 indicated a monocytic immune response and activation of macrophages [19]. Complement and coagulation factors remained on baseline level, as inferred from the mildest subtype 3, which was interpreted as the sepsis plasma proteome baseline of the patients in the investigated cohort. This assumption was supported for example by VCAM1 and ICAM1, which at day 1 showed the lowest levels in cluster 3 (regulation pattern α), and for which increasing plasma levels are well known to be related to sepsis severity [20].

Although we found SOFA scores to be gradually decreasing between clusters 1, 2 and 3, no significant difference in 30-day mortality was observed. Contrary to these findings, the ALBIOS trial reported higher mortality after 90 days to be associated with increased IgG and IgA levels [21]. In addition, a decrease of high-density lipoproteins has been reported to be predictive for sepsis mortality [22].

Both observations match the increased SOFA scores and cytokine levels in cluster 1, which themselves are predictive for higher mortality [23], so a worse outcome for patients in cluster 1 might have been anticipated. Unfortunately, a later endpoint such as 90-day survival and adverse events like secondary infections, were not assessed in our study.

Sepsis subtypes, also termed subclasses, phenotypes or endotypes, often used synonymously [24], are considered a major way towards personalized therapy in sepsis. In contrast to clinical subtypes which rely mostly on clinical routine and laboratory data [13, 25], molecular subtypes have the potential to provide direct insights into pathophysiology. Until now, multiple molecular subtypes have been reported and were often based on transcriptome analysis using whole blood or immune cells [14, 26–28], but also multiplex cytokine measurements or the combination of both [29, 30].

Recently, the first study describing three molecular subtypes based on plasma proteomics data has been published [10]. Of note, Mi and colleagues also observed a subtype which was characterized by high Ig levels and low levels of apolipoproteins as well complement activation (termed SPC1). The overlap with our observations implies generalizability of this subtype, and highlights its potential clinical relevance, for example in patient stratification in clinical trials. One therapeutic consideration that obviously may be influenced by plasma proteome subtypes is the use of intravenous (IgM-enriched) immunoglobulin preparations, which have shown heterogenous results in clinical trials and are still under debate [31, 32].

While sepsis plasma proteome subtypes provide clinically relevant information, the assignment of new patients to the subtypes remains challenging. As mass spectrometry-based plasma proteomics are currently unlikely to be implemented into clinical routine, we generated a ML classifier based on only 10 proteins. Additional Ig quantities were measured on a routine diagnostics platform and assays for albumin and antithrombin III (SERPINC1) are readily available. In future, the remaining proteins could be measured using immunoassays, ideally in a multiplex manner. The classifier differentiates only subtypes 1-3, as cluster 0 was too small to generate a robust ML model. However, patients with most severe sepsis might be identified based on clinical routine data as described previously [33]. The most important protein for differentiation of plasma subtypes 1 – 3 was Complement C9 (C9). C9 is a component of the membrane attack complex (MAC) and low levels of C9 were found to be associated with septic shock in patients with Gam-negative bacteremia [34].

Complement factor C2, part of the classical pathway, was found to be the second most important protein, underlining the relevance of the complement system in sepsis. Another highly relevant feature was serum albumin (ALB). Interestingly, low levels of albumin, as indicative for cluster 1, were previously reported to be associated with poor prognosis [35, 36], which supports the impression that cluster 1 might be associated with a worse outcome than observed using the 30-day survival endpoint. In addition, a decrease of the kallikrein inhibitor kallistatin (SERPINA4), also indicative for cluster 1, was reported to be associated with septic shock and sepsis mortality [37].

Antithrombin III (SERPINC1) as well as Alpha-2-antiplasmin (SERPINF2) are both important regulators of the coagulation cascade and were reported to be associated with sepsis mortality and organ dysfunction [7]. The alpha-trypsin inhibitor ITIH3 has been described in sepsis [38] as well as to be related to mortality in severe COVID-19 [39] and the activity of the serum paraoxonase PON1 has been shown to be predictive for 30-day mortality [40]. Taken together, there is ample evidence for the diagnostic relevance of most of the selected proteins, underlining the value of the model and suggesting that of our findings might be applicable in independent cohorts. The developed model illustrates that the transfer between different technological platforms is not necessary an obstacle to transferring proteomics subtypes into the clinics and opens up a perspective on how patient stratification could be realized.

### Conclusions

The identified sepsis plasma proteome subtypes are characterized by distinct features that provide insights into the underlying disease mechanisms and immune processes of this heterogeneous syndrome. These findings allow conclusions on benefits and disadvantages of therapeutic interventions, and have the potential to support the predictive enrichment of patients in clinical trials. In a clinical setting, patient stratification could be achieved by measuring a representative biomarker panel. These findings represent a significant advancement towards precision medicine in sepsis management.

## Abbreviations

AKI: Acute kidney injury
ALT: Alanine transaminase
AST: Aspartate transaminase
AP: Alkaline phosphatase
aPTT: Activated partial thromboplastin time
AUROC: Area under the receiver operating characteristic curve
CHE: Cholinesterase
CNS: Central nervous system
CRP: C-reactive protein
DIC: Disseminated intravascular coagulation
FDR: False discovery rate
GO: Gene ontology
ICU: Intensive care unit
Ig: Immunoglobulin
INR: International normalized ratio
IQR: Interquartile range
LDH: Lactate dehydrogenase
LOD: Limit of detection
MCCV: Monte carlo cross-validation
ML: Machine learning
PCA: Principal component analysis
SHAP: SHapley additive exPlanations
SOFA: Sepsis-related organ failure assessment score

## Appendix

### Data availability

The mass spectrometry proteomics data have been deposited to the ProteomeXchange Consortium via the PRIDE partner repository with the dataset identifier PXD064125.

### Ethics approval and consent to participate

The SepsisDataNet.NRW and CovidDataNet.NRW studies were approved by the Ethics Committee of the Medical Faculty of Ruhr-University Bochum (Registration No. 5047–14 and 19-6606_6-BR, respectively) or the responsible ethics committee of each respective study center and conducted in accordance with the revised Declaration of Helsinki. Written informed consent was obtained from the patients or a legal representative.

## Acknowledgement

The authors thank the SepsisDataNet.NRW and CovidDataNet.NRW research groups. The authors thank Kristin Fuchs and Birgit Zülch for excellent technical assistance.

## Author contributions

Conceptualization: TB, MA, BS, HN, Writing the original draft: TB, KK, HN, Revision of original draft: MB, FG, KS, LP, MU, TR, BK, KR, DZ, DH, TG, FSE, ME, SP, NB, AZ, KM, BS, MA, Data generation & patient recruitment: MB, FG, MBP, TR, MU, LB, BK, KR, TG, DZ, UL, DH, SFE, Data analysis: TB, KK, KS, HN, Supervision: MA, BS

## References

1. Singer M, Deutschman CS, Seymour CW, Shankar-Hari M, Annane D, Bauer M, Bellomo R, Bernard GR, Chiche JD, Coopersmith CM et al: The Third International Consensus Definitions for Sepsis and Septic Shock (Sepsis-3). Jama 2016, 315(8):801–810.

2. Rudd KE, Johnson SC, Agesa KM, Shackelford KA, Tsoi D, Kievlan DR, Colombara DV, Ikuta KS, Kissoon N, Finfer S et al: Global, regional, and national sepsis incidence and mortality, 1990-2017: analysis for the Global Burden of Disease Study. Lancet 2020, 395(10219):200–211.

3. Evans L, Rhodes A, Alhazzani W, Antonelli M, Coopersmith CM, French C, Machado FR, McIntyre L, Ostermann M, Prescott HC et al: Surviving sepsis campaign: international guidelines for management of sepsis and septic shock 2021. Intensive care medicine 2021, 47(11):1181–1247.

4. Marshall JC: Why have clinical trials in sepsis failed? Trends in molecular medicine 2014, 20(4):195–203.

5. Scicluna BP, Baillie JK: The Search for Efficacious New Therapies in Sepsis Needs to Embrace Heterogeneity. American journal of respiratory and critical care medicine 2019, 199(8):936–938.

6. Palmowski L, Weber M, Bayer M, Mi Y, Schork K, Eisenacher M, Nowak H, Rahmel T, Bergmann L, Witowski A et al: Mortality-associated plasma proteome dynamics in a prospective multicentre sepsis cohort. EBioMedicine 2025, 111:105508.

7. Ruiz-Sanmartin A, Ribas V, Sunol D, Chiscano-Camon L, Palmada C, Bajana I, Larrosa N, Gonzalez JJ, Canela N, Ferrer R et al: Characterization of a proteomic profile associated with organ dysfunction and mortality of sepsis and septic shock. PloS one 2022, 17(12):e0278708.

8. Pimienta G, Heithoff DM, Rosa-Campos A, Tran M, Esko JD, Mahan MJ, Marth JD, Smith JW: Plasma Proteome Signature of Sepsis: a Functionally Connected Protein Network. Proteomics 2019, 19(5):e1800389.

9. Liang X, Wu T, Chen Q, Jiang J, Jiang Y, Ruan Y, Zhang H, Zhang S, Zhang C, Chen P et al: Serum proteomics reveals disorder of lipoprotein metabolism in sepsis. Life science alliance 2021, 4(10).

10. Mi Y, Burnham KL, Charles PD, Heilig R, Vendrell I, Whalley J, Torrance HD, Antcliffe DB, May SM, Neville MJ et al: High-throughput mass spectrometry maps the sepsis plasma proteome and differences in patient response. Science translational medicine 2024, 16(750):eadh0185.

11. Seymour CW, Kennedy JN, Wang S, Chang CH, Elliott CF, Xu Z, Berry S, Clermont G, Cooper G, Gomez H et al: Derivation, Validation, and Potential Treatment Implications of Novel Clinical Phenotypes for Sepsis. Jama 2019, 321(20):2003–2017.

12. Chenoweth JG, Brandsma J, Striegel DA, Genzor P, Chiyka E, Blair PW, Krishnan S, Dogbe E, Boakye I, Fogel GB et al: Sepsis endotypes identified by host gene expression across global cohorts. Communications medicine 2024, 4(1):120.

13. Xu Z, Mao C, Su C, Zhang H, Siempos I, Torres LK, Pan D, Luo Y, Schenck EJ, Wang F: Sepsis subphenotyping based on organ dysfunction trajectory. Critical care 2022, 26(1):197.

14. Davenport EE, Burnham KL, Radhakrishnan J, Humburg P, Hutton P, Mills TC, Rautanen A, Gordon AC, Garrard C, Hill AV et al: Genomic landscape of the individual host response and outcomes in sepsis: a prospective cohort study. The Lancet Respiratory medicine 2016, 4(4):259–271.

15. van Amstel RBE, Kennedy JN, Scicluna BP, Bos LDJ, Peters-Sengers H, Butler JM, Cano-Gamez E, Knight JC, Vlaar APJ, Cremer OL et al: Uncovering heterogeneity in sepsis: a comparative analysis of subphenotypes. Intensive care medicine 2023, 49(11):1360–1369.

16. Zhang Z, Chen L, Liu X, Yang J, Huang J, Yang Q, Hu Q, Jin K, Celi LA, Hong Y: Exploring disease axes as an alternative to distinct clusters for characterizing sepsis heterogeneity. Intensive care medicine 2023, 49(11):1349–1359.

17. Bracht T, Kleefisch D, Schork K, Witzke KE, Chen W, Bayer M, Hovanec J, Johnen G, Meier S, Ko YD et al: Plasma Proteomics Enable Differentiation of Lung Adenocarcinoma from Chronic Obstructive Pulmonary Disease (COPD). International journal of molecular sciences 2022, 23(19).

18. Unterberg M, Ehrentraut SF, Bracht T, Wolf A, Haberl H, von Busch A, Rump K, Ziehe D, Bazzi M, Thon P et al: Human cytomegalovirus seropositivity is associated with reduced patient survival during sepsis. Critical care 2023, 27(1):417.

19. Reissing J, Lutz P, Frissen M, Ibidapo-Obe O, Reuken PA, Wirtz TH, Stengel S, Quickert S, Rooney M, Grosse K et al: Immunomodulatory receptor VSIG4 is released during spontaneous bacterial peritonitis and predicts short-term mortality. JHEP reports: innovation in hepatology 2022, 4(1):100391.

20. He J, Duan M, Zhuang H: ICAM1 and VCAM1 are associated with outcome in patients with sepsis: A systematic review and meta-analysis. Heliyon 2024, 10(21):e40003.

21. Alagna L, Meessen J, Bellani G, Albiero D, Caironi P, Principale I, Vivona L, Grasselli G, Motta F, Agnelli NM et al: Higher levels of IgA and IgG at sepsis onset are associated with higher mortality: results from the Albumin Italian Outcome Sepsis (ALBIOS) trial. Annals of intensive care 2021, 11(1):161.

22. Chien JY, Jerng JS, Yu CJ, Yang PC: Low serum level of high-density lipoprotein cholesterol is a poor prognostic factor for severe sepsis. Critical care medicine 2005, 33(8):1688–1693.

23. Ferreira FL, Bota DP, Bross A, Melot C, Vincent JL: Serial evaluation of the SOFA score to predict outcome in critically ill patients. Jama 2001, 286(14):1754–1758.

24. DeMerle KM, Angus DC, Baillie JK, Brant E, Calfee CS, Carcillo J, Chang CH, Dickson R, Evans I, Gordon AC et al: Sepsis Subclasses: A Framework for Development and Interpretation. Critical care medicine 2021, 49(5):748–759.

25. Gardlund B, Dmitrieva NO, Pieper CF, Finfer S, Marshall JC, Taylor Thompson B: Six subphenotypes in septic shock: Latent class analysis of the PROWESS Shock study. Journal of critical care 2018, 47:70–79.

26. Sweeney TE, Azad TD, Donato M, Haynes WA, Perumal TM, Henao R, Bermejo-Martin JF, Almansa R, Tamayo E, Howrylak JA et al: Unsupervised Analysis of Transcriptomics in Bacterial Sepsis Across Multiple Datasets Reveals Three Robust Clusters. Critical care medicine 2018, 46(6):915–925.

27. Scicluna BP, van Vught LA, Zwinderman AH, Wiewel MA, Davenport EE, Burnham KL, Nurnberg P, Schultz MJ, Horn J, Cremer OL et al: Classification of patients with sepsis according to blood genomic endotype: a prospective cohort study. The Lancet Respiratory medicine 2017, 5(10):816–826.

28. Chenoweth JG, Colantuoni C, Striegel DA, Genzor P, Brandsma J, Blair PW, Krishnan S, Chiyka E, Fazli M, Mehta R et al: Gene expression signatures in blood from a West African sepsis cohort define host response phenotypes. Nature communications 2024, 15(1):4606.

29. Antcliffe DB, Mi Y, Santhakumaran S, Burnham KL, Prevost AT, Ward JK, Marshall TJ, Bradley C, Al-Beidh F, Hutton P et al: Patient stratification using plasma cytokines and their regulators in sepsis: relationship to outcomes, treatment effect and leucocyte transcriptomic subphenotypes. Thorax 2024, 79(6):515–523.

30. Bodinier M, Peronnet E, Llitjos JF, Kreitmann L, Brengel-Pesce K, Rimmele T, Fleurie A, Textoris J, Venet F, Maucort-Boulch D et al: Integrated clustering of multiple immune marker trajectories reveals different immunotypes in severely injured patients. Critical care 2024, 28(1):240.

31. Soares MO, Welton NJ, Harrison DA, Peura P, Shankar-Hari M, Harvey SE, Madan J, Ades AE, Rowan KM, Palmer SJ: Intravenous immunoglobulin for severe sepsis and septic shock: clinical effectiveness, cost-effectiveness and value of a further randomised controlled trial. Critical care 2014, 18(6):649.

32. Cui J, Wei X, Lv H, Li Y, Li P, Chen Z, Liu G: The clinical efficacy of intravenous IgM-enriched immunoglobulin (pentaglobin) in sepsis or septic shock: a meta-analysis with trial sequential analysis. Annals of intensive care 2019, 9(1):27.

33. Bracht T, Weber M, Kappler K, Palmowski L, Bayer M, Schork K, Rahmel T, Unterberg M, Haberl H, Wolf A et al: Machine learning identifies clinical sepsis phenotypes that translate to the plasma proteome: a prospective cohort study. medRxiv 2025:2025.2004.2011.25325574.

34. Eichenberger EM, Dagher M, Ruffin F, Park L, Hersh L, Sivapalasingam S, Fowler VG, Jr., Prasad BC: Complement levels in patients with bloodstream infection due to Staphylococcus aureus or Gram-negative bacteria. European journal of clinical microbiology & infectious diseases: official publication of the European Society of Clinical Microbiology 2020, 39(11):2121–2131.

35. Artero A, Zaragoza R, Camarena JJ, Sancho S, Gonzalez R, Nogueira JM: Prognostic factors of mortality in patients with community-acquired bloodstream infection with severe sepsis and septic shock. Journal of critical care2010, 25(2):276–281.

36. Yin M, Si L, Qin W, Li C, Zhang J, Yang H, Han H, Zhang F, Ding S, Zhou M et al: Predictive Value of Serum Albumin Level for the Prognosis of Severe Sepsis Without Exogenous Human Albumin Administration: A Prospective Cohort Study. Journal of intensive care medicine 2018, 33(12):687–694.

37. Lin WC, Chen CW, Chao L, Chao J, Lin YS: Plasma kallistatin in critically ill patients with severe sepsis and septic shock. PloS one 2017, 12(5):e0178387.

38. Thavarajah T, Dos Santos CC, Slutsky AS, Marshall JC, Bowden P, Romaschin A, Marshall JG: The plasma peptides of sepsis. Clinical proteomics 2020, 17:26.

39. Vollmy F, van den Toorn H, Zenezini Chiozzi R, Zucchetti O, Papi A, Volta CA, Marracino L, Vieceli Dalla Sega F, Fortini F, Demichev V et al: A serum proteome signature to predict mortality in severe COVID-19 patients. Life science alliance 2021, 4(9).

40. Shukeri W, Ralib AM, Abdulah NZ, Mat-Nor MB: Sepsis mortality score for the prediction of mortality in septic patients. Journal of critical care 2018, 43:163–168.

