## Supplementary Material for "Plasma proteomics identifies molecular subtypes in sepsis"

**Table of content**

| **#** | **Title** |
| --- | --- |
| **1** | Supplementary Table 1: Protein features that were considered for machine learning. |
| **2** | Supplementary Table 2: Correlating feature pairs in the dataset […] |
| **3** | Supplementary Table 4: Clinical features that were tested for feature importance in machine learning. |
| **4** | Supplementary Table 5: Hyperparameters for the random forest models for the prediction with clinical and proteomics data. |
| **5** | Supplementary Table 7: Protein regulation profiles. |
| **6** | Supplementary Figure 1: Quality control for proteomics data normalization. |
| **7** | Supplementary Figure 2: Silhouette curves that were used to determine of the optimal number of clusters. |
| **8** | Supplementary Figure 3: Framework for machine learning and feature selection. |
| **9** | Supplementary Figure 4: Time of ICU stay and hospital stay. |
| **10** | Supplementary Figure 5: Clinical characteristics of the molecular subtypes. |
| **11** | Supplementary Figure 6: Proteome characteristics of plasma proteome subtypes at day 4. |
| **12** | Supplementary Figure 7: Identified protein abundance patterns for day 1 and day 4. |
| **13** | Supplementary Figure 8: Feature Selection for the machine learning classifier. |
| **14** | Supplementary Figure 9: SHAP summary plots for the machine learning classifier. |

**Supplementary Table 1:** Protein features that were considered for machine learning.

| # | Protein | Gene | Protein Description |
| --- | --- | --- | --- |
| 1 | O00187 | MASP2 | Mannan-binding lectin serine protease 2 |
| 2 | O00391 | QSOX1 | Sulfhydryl oxidase 1 |
| 3 | O14791 | APOL1 | Apolipoprotein L1 |
| 4 | O43866 | CD5L | CD5 antigen-like |
| 5 | O75636 | FCN3 | Ficolin-3 |
| 6 | O75882 | ATRN | Attractin |
| 7 | O95445 | APOM | Apolipoprotein M |
| 8 | P00338 | LDHA | L-lactate dehydrogenase A chain |
| 9 | P00450 | CP | Ceruloplasmin |
| 10 | P00488 | F13A1 | Coagulation factor XIII A chain |
| 11 | P00734 | F2 | Prothrombin |
| 12 | P00736 | C1R | Complement C1r subcomponent |
| 13 | P00738 | HP | Haptoglobin |
| 14 | P00739 | HPR | Haptoglobin-related protein |
| 15 | P00740 | F9 | Coagulation factor IX |
| 16 | P00742 | F10 | Coagulation factor X |
| 17 | P00747 | PLG | Plasminogen |
| 18 | P00748 | F12 | Coagulation factor XII |
| 19 | P00751 | CFB | Complement factor B |
| 20 | P01008 | SERPINC1 | Antithrombin-III |
| 21 | P01009 | SERPINA1 | Alpha-1-antitrypsin |
| 22 | P01011 | SERPINA3 | Alpha-1-antichymotrypsin |
| 23 | P01019 | AGT | Angiotensinogen |
| 24 | P01023 | A2M | Alpha-2-macroglobulin |
| 25 | P01024 | C3 | Complement C3 |
| 26 | P01031 | C5 | Complement C5 |
| 27 | P01034 | CST3 | Cystatin-C |
| 28 | P01042 | KNG1 | Kininogen-1 |
| 29 | P02144 | MB | Myoglobin |
| 30 | P02647 | APOA1 | Apolipoprotein A-I |
| 31 | P02649 | APOE | Apolipoprotein E |
| 32 | P02652 | APOA2 | Apolipoprotein A-II |
| 33 | P02654 | APOC1 | Apolipoprotein C-I |
| 34 | P02656 | APOC3 | Apolipoprotein C-III |
| 35 | P02743 | APCS | Serum amyloid P-component |
| 36 | P02747 | C1QC | Complement C1q subcomponent subunit C |
| 37 | P02748 | C9 | Complement component C9 |
| 38 | P02749 | APOH | Beta-2-glycoprotein 1 |
| 39 | P02750 | LRG1 | Leucine-rich alpha-2-glycoprotein |
| 40 | P02751 | FN1 | Fibronectin |
| 41 | P02753 | RBP4 | Retinol-binding protein 4 |
| 42 | P02760 | AMBP | Protein AMBP |
| 43 | P02763 | ORM1 | Alpha-1-acid glycoprotein 1 |
| 44 | P02765 | AHSG | Alpha-2-HS-glycoprotein |
| 45 | P02766 | TTR | Transthyretin |
| 46 | P02768 | ALB | Albumin |
| 47 | P02774 | GC | Vitamin D-binding protein |
| 48 | P02775 | PPBP | Platelet basic protein |
| 49 | P02786 | TFRC | Transferrin receptor protein 1 |
| 50 | P02787 | TF | Serotransferrin |
| 51 | P02790 | HPX | Hemopexin |
| 52 | P03951 | F11 | Coagulation factor XI |
| 53 | P03952 | KLKB1 | Plasma kallikrein |
| 54 | P04003 | C4BPA | C4b-binding protein alpha chain |
| 55 | P04004 | VTN | Vitronectin |
| 56 | P04040 | CAT | Catalase |
| 57 | P04070 | PROC | Vitamin K-dependent protein C |
| 58 | P04075 | ALDOA | Fructose-bisphosphate aldolase A |
| 59 | P04114 | APOB | Apolipoprotein B-100 |
| 60 | P04180 | LCAT | Phosphatidylcholine-sterol acyltransferase |
| 61 | P04196 | HRG | Histidine-rich glycoprotein |
| 62 | P04217 | A1BG | Alpha-1B-glycoprotein |
| 63 | P04264 | KRT1 | Keratin, type II cytoskeletal 1 |
| 64 | P04275 | VWF | von Willebrand factor |
| 65 | P04278 | SHBG | Sex hormone-binding globulin |
| 66 | P04406 | GAPDH | Glyceraldehyde-3-phosphate dehydrogenase |
| 67 | P05062 | ALDOB | Fructose-bisphosphate aldolase B |
| 68 | P05090 | APOD | Apolipoprotein D |
| 69 | P05109 | S100A8 | Protein S100-A8 |
| 70 | P05154 | SERPINA5 | Plasma serine protease inhibitor |
| 71 | P05155 | SERPING1 | Plasma protease C1 inhibitor |
| 72 | P05156 | CFI | Complement factor I |
| 73 | P05160 | F13B | Coagulation factor XIII B chain |
| 74 | P05452 | CLEC3B | Tetranectin |
| 75 | P05543 | SERPINA7 | Thyroxine-binding globulin |
| 76 | P05546 | SERPIND1 | Heparin cofactor 2 |
| 77 | P06396 | GSN | Gelsolin |
| 78 | P06681 | C2 | Complement C2 |
| 79 | P06727 | APOA4 | Apolipoprotein A-IV |
| 80 | P07195 | LDHB | L-lactate dehydrogenase B chain |
| 81 | P07225 | PROS1 | Vitamin K-dependent protein S |
| 82 | P07357 | C8A | Complement component C8 alpha chain |
| 83 | P07358 | C8B | Complement component C8 beta chain |
| 84 | P07359 | GP1BA | Platelet glycoprotein Ib alpha chain |
| 85 | P07360 | C8G | Complement component C8 gamma chain |
| 86 | P08185 | SERPINA6 | Corticosteroid-binding globulin |
| 87 | P08294 | SOD3 | Extracellular superoxide dismutase [Cu-Zn] |
| 88 | P08519 | LPA | Apolipoprotein(a) |
| 89 | P08571 | CD14 | Monocyte differentiation antigen CD14 |
| 90 | P08603 | CFH | Complement factor H |
| 91 | P08697 | SERPINF2 | Alpha-2-antiplasmin |
| 92 | P0C0L4 | C4A | Complement C4-A |
| 93 | P0C0L5 | C4B_2 | Complement C4-B |
| 94 | P0C0S8 | H2AC17 | Histone H2A type 1 |
| 95 | P0DJI8 | SAA1 | Serum amyloid A-1 protein |
| 96 | P10643 | C7 | Complement component C7 |
| 97 | P10909 | CLU | Clusterin |
| 98 | P11021 | HSPA5 | Endoplasmic reticulum chaperone BiP |
| 99 | P11226 | MBL2 | Mannose-binding protein C |
| 100 | P12259 | F5 | Coagulation factor V |
| 101 | P13671 | C6 | Complement component C6 |
| 102 | P13796 | LCP1 | Plastin-2 |
| 103 | P14151 | SELL | L-selectin |
| 104 | P18065 | IGFBP2 | Insulin-like growth factor-binding protein 2 |
| 105 | P18206 | VCL | Vinculin |
| 106 | P18428 | LBP | Lipopolysaccharide-binding protein |
| 107 | P19320 | VCAM1 | Vascular cell adhesion protein 1 |
| 108 | P19652 | ORM2 | Alpha-1-acid glycoprotein 2 |
| 109 | P19823 | ITIH2 | Inter-alpha-trypsin inhibitor heavy chain H2 |
| 110 | P20742 | PZP | Pregnancy zone protein |
| 111 | P20851 | C4BPB | C4b-binding protein beta chain |
| 112 | P22352 | GPX3 | Glutathione peroxidase 3 |
| 113 | P22792 | CPN2 | Carboxypeptidase N subunit 2 |
| 114 | P22891 | PROZ | Vitamin K-dependent protein Z |
| 115 | P23142 | FBLN1 | Fibulin-1 |
| 116 | P25311 | AZGP1 | Zinc-alpha-2-glycoprotein |
| 117 | P26927 | MST1 | Hepatocyte growth factor-like protein |
| 118 | P27169 | PON1 | Serum paraoxonase/arylesterase 1 |
| 119 | P29622 | SERPINA4 | Kallistatin |
| 120 | P32119 | PRDX2 | Peroxiredoxin-2 |
| 121 | P33908 | MAN1A1 | Mannosyl-oligosaccharide 1,2-alpha-mannosidase IA |
| 122 | P35542 | SAA4 | Serum amyloid A-4 protein |
| 123 | P35858 | IGFALS | Insulin-like growth factor-binding protein complex acid labile subunit |
| 124 | P36955 | SERPINF1 | Pigment epithelium-derived factor |
| 125 | P43251 | BTD | Biotinidase |
| 126 | P43652 | AFM | Afamin |
| 127 | P48740 | MASP1 | Mannan-binding lectin serine protease 1 |
| 128 | P49908 | SELENOP | Selenoprotein P |
| 129 | P51884 | LUM | Lumican |
| 130 | P55056 | APOC4 | Apolipoprotein C-IV |
| 131 | P55058 | PLTP | Phospholipid transfer protein |
| 132 | P60709 | ACTB | Actin, cytoplasmic 1 |
| 133 | P61626 | LYZ | Lysozyme C |
| 134 | P61769 | B2M | Beta-2-microglobulin |
| 135 | P78417 | GSTO1 | Glutathione S-transferase omega-1 |
| 136 | P80108 | GPLD1 | Phosphatidylinositol-glycan-specific phospholipase D |
| 137 | Q03591 | CFHR1 | Complement factor H-related protein 1 |
| 138 | Q04756 | HGFAC | Hepatocyte growth factor activator |
| 139 | Q06033 | ITIH3 | Inter-alpha-trypsin inhibitor heavy chain H3 |
| 140 | Q08380 | LGALS3BP | Galectin-3-binding protein |
| 141 | Q08830 | FGL1 | Fibrinogen-like protein 1 |
| 142 | Q12805 | EFEMP1 | EGF-containing fibulin-like extracellular matrix protein 1 |
| 143 | Q13790 | APOF | Apolipoprotein F |
| 144 | Q14520 | HABP2 | Hyaluronan-binding protein 2 |
| 145 | Q14624 | ITIH4 | Inter-alpha-trypsin inhibitor heavy chain H4 |
| 146 | Q15582 | TGFBI | Transforming growth factor-beta-induced protein ig-h3 |
| 147 | Q16610 | ECM1 | Extracellular matrix protein 1 |
| 148 | Q92496 | CFHR4 | Complement factor H-related protein 4 |
| 149 | Q92954 | PRG4 | Proteoglycan 4 |
| 150 | Q96IY4 | CPB2 | Carboxypeptidase B2 |
| 151 | Q96PD5 | PGLYRP2 | N-acetylmuramoyl-L-alanine amidase |
| 152 | Q9BXR6 | CFHR5 | Complement factor H-related protein 5 |
| 153 | Q9NZP8 | C1RL | Complement C1r subcomponent-like protein |
| 154 | Q9UGM5 | FETUB | Fetuin-B |
| 155 | Q9UK55 | SERPINA10 | Protein Z-dependent protease inhibitor |
| 156 | Q9Y6R7 | FCGBP | IgGFc-binding protein |
| 157 | O43286 | B4GALT5 | Beta-1,4-galactosyltransferase 5 |
| 158 | P07339 | CTSD | Cathepsin D |
| 159 | P49913 | CAMP | Cathelicidin antimicrobial peptide |
| 160 | Q68DL7 | C18orf63 | Uncharacterized protein C18orf63 |
| 161 | Q9NP80 | PNPLA8 | Calcium-independent phospholipase A2-gamma |

**Supplementary Table 2:** Correlating feature pairs in the dataset as evaluated by the Pearson Correlation Coefficient.

| Feature 1 | Feature 2 | Correlation |
| --- | --- | --- |
| BE_MEDIAN* | HCO3-_MAX | 0.91 |
| AST_MAX | ALT_MAX | 0.80 |
| AST_MAX | LDH_MAX | 0.82 |
| ALT_MAX | LDH_MAX | 0.87 |
| INR_MAX* | QUICK_MIN | -0.72 |
| BP_dia_MEDIAN | BP_mean_MEDIAN* | 0.87 |
| BP_mean_MEDIAN* | BP_sys_MEDIAN | 0.72 |
| pO2_MIN | sO2_MIN* | 0.83 |
| H2BC12 | H2AC17* | 0.92 |
| H2BC12 | TKT | 0.70 |
| LDHA* | GAPDH | 0.72 |
| LDHA* | HSP90AA1 | 0.72 |
| C1R* | C1S | 0.78 |
| CFD | B2M* | 0.74 |
| CA1 | PRDX2* | 0.79 |
| CA1 | CA2 | 0.70 |
| SERPINA1 | SERPINA3* | 0.71 |
| APOC2 | APOC3* | 0.74 |
| C1QA | C1QB* | 0.85 |
| C1QA | C1QC* | 0.87 |
| C1QB | C1QC* | 0.87 |
| ORM1* | ORM2 | 0.74 |
| PPBP* | PF4 | 0.85 |
| PPBP | THBS1 | 0.76 |
| PF4 | THBS1 | 0.79 |
| PF4 | FLNA | 0.71 |
| KRT1* | KRT10 | 0.87 |
| KRT1* | KRT9 | 0.88 |
| KRT1* | KRT2 | 0.90 |
| S100A8* | S100A9 | 0.92 |
| SAA1* | SAA2 | 0.81 |
| KRT10 | KRT9 | 0.73 |
| KRT10 | KRT2 | 0.94 |
| CPN1 | CPN2* | 0.75 |
| IGFBP3 | IGFALS* | 0.72 |
| VCL* | FLNA | 0.75 |
| VCL* | TLN1 | 0.75 |
| ITIH2* | ITIH1 | 0.85 |
| FLNA | TLN1 | 0.87 |
| PRDX2* | CA2 | 0.75 |
| KRT9 | KRT2 | 0.73 |
| ACTB* | ACTC1 | 0.80 |

*Features marked with a star (*) remained in the dataset, the other features were dropped.*

**Supplementary Table 4:** Clinical features that were tested for feature importance in machine learning.

| # | Abbreviation | Description | Location parameter |
| --- | --- | --- | --- |
| 1 | AP_MAX | alkaline phosphatase level (serum) | maximum |
| 2 | BE_MEDIAN | base excess (arterial blood gas) | median |
| 3 | BR_total_MAX | total bilirubin level (serum) | maximum |
| 4 | CK_MAX | creatine kinase level (serum) | maximum |
| 5 | CRP_MAX | C-reactive protein level (serum) | maximum |
| 6 | CA_MIN | ionized calcium level (serum or point-of-care testing) | minimum |
| 7 | TC_MAX | total cholesterol level (serum) | maximum |
| 8 | PROT_total_MIN | total protein level (serum) | minimum |
| 9 | FIBR_MIN | fibrinogen level (serum) | minimum |
| 10 | AST_MAX | aspartate transaminase level (serum) | maximum |
| 11 | ALT_MAX | alanine transaminase level (serum) | maximum |
| 12 | HF_MEDIAN | heart rate | median |
| 13 | UREA_N_MAX | urea nitrogen level (serum) | maximum |
| 14 | HB_MIN | hemoglobin level (blood) | minimum |
| 15 | BP_mean_MEDIAN | mean arterial blood pressure | median |
| 16 | INR_MAX | international normalized ratio of thrombin time | maximum |
| 17 | K_MAX | potassium level (serum) | maximum |
| 18 | CREA_MAX | creatinine level (serum) | maximum |
| 19 | LDH_MAX | lactate dehydrogenase level (serum) | maximum |
| 20 | PCT_MAX | procalcitonin level (serum) | maximum |
| 21 | PHOS_MIN | phosphate level (serum) | minimum |
| 22 | SpO2_MIN | peripheral oxygen saturation | minimum |
| 23 | TEMP_MAX | body temperature | maximum |
| 24 | PLT_MIN | platelet count (blood) | minimum |
| 25 | APTT_MAX | activated partial thromboplastin time | maximum |
| 26 | pH_MIN | pH (potential of hydrogen, arterial blood gas) | minimum |
| 27 | LAC_MAX | lactate level (point-of-care testing) | maximum |
| 28 | IgA | Immunoglobulin A | - |
| 29 | IgG | Immunoglobulin G | - |
| 30 | IgM | Immunoglobulin M | - |

**Supplementary Table 5:** Hyperparameters for the random forest models for the prediction with clinical and proteomics data.

| **Hyperparameter** | **Chosen value** |
| --- | --- |
| n_estimators | 100 |
| min_samples_leaf | 5 |
| min_samples_split | 5 |
| max_depth | 10 |
| Class_weight | “balanced” |

**Supplementary Table 7:** Protein regulation profiles. Uniprot identifier and Gene names given for the regulation patterns that were distinguished according to hierarchical cluster analysis (Figure 2a, Supplementary Figure 2a).

| **Day 1** | | | | | | | | | |
| --- | --- | --- | --- | --- | --- | --- | --- | --- | --- |
| **α pattern** | | **β pattern** | | **γ pattern** | | **δ pattern** | | **ε pattern** | |
| **Identifier** | **Gene** | **Identifier** | **Gene** | **Identifier** | **Gene** | **Identifier** | **Gene** | **Identifier** | **Gene** |
| A0A0B4J1U3 | IGLV1-36 | O75882 | ATRN | O00187 | MASP2 | O60814 | H2BC12 | A0A075B6H7 | IGKV3-7 |
| P01034 | CST3 | P00488 | F13A1 | P00738 | HP | O75874 | IDH1 | A0A075B6I0 | IGLV8-61 |
| P01833 | PIGR | P00739 | HPR | P00740 | F9 | P00325 | ADH1B | A0A075B6K4 | IGLV3-10 |
| P04264 | KRT1 | P01024 | C3 | P02741 | CRP | P00338 | LDHA | A0A075B6K5 | IGLV3-9 |
| P04275 | VWF | P02647 | APOA1 | P02743 | APCS | P00352 | ALDH1A1 | A0A075B6P5 | IGKV2-28 |
| P05362 | ICAM1 | P02654 | APOC1 | P02748 | C9 | P00915 | CA1 | A0A075B6S2 | IGKV2D-29 |
| P06727 | APOA4 | P02655 | APOC2 | P02775 | PPBP | P02144 | MB | A0A075B6S5 | IGKV1-27 |
| P18065 | IGFBP2 | P02656 | APOC3 | P03951 | F11 | P04040 | CAT | A0A087WSY6 | IGKV3D-15 |
| P19320 | VCAM1 | P02753 | RBP4 | P04004 | VTN | P04075 | ALDOA | A0A087WSZ0 | IGKV1D-8 |
| P24158 | PRTN3 | P02766 | TTR | P07357 | C8A | P04179 | SOD2 | A0A0A0MRZ8 | IGKV3D-11 |
| P35527 | KRT9 | P02768 | ALB | P07358 | C8B | P04406 | GAPDH | A0A0A0MS15 | IGHV3-49 |
| P61626 | LYZ | P04070 | PROC | P08514 | ITGA2B | P05062 | ALDOB | A0A0B4J1U7 | IGHV6-1 |
| Q15582 | TGFBI | P04180 | LCAT | P0DJI8 | SAA1 | P05109 | S100A8 | A0A0B4J1V0 | IGHV3-15 |
| Q9Y6R7 | FCGBP | P04196 | HRG | P0DJI9 | SAA2 | P06702 | S100A9 | A0A0B4J1V2 | IGHV2-26 |
| O43286 | B4GALT5 | P04278 | SHBG | P11226 | MBL2 | P06733 | ENO1 | A0A0B4J1X5 | IGHV3-74 |
| O75019 | LILRA1 | P05154 | SERPINA5 | P12259 | F5 | P07237 | P4HB | A0A0B4J1Y8 | IGLV9-49 |
| Q9Y279 | VSIG4 | P05156 | CFI | P13671 | C6 | P07900 | HSP90AA1 | A0A0B4J1Y9 | IGHV3-72 |
| Q9Y5Y7 | LYVE1 | P05160 | F13B | P14151 | SELL | P08238 | HSP90AB1 | A0A0B4J2D9 | IGKV1D-13 |
|  |  | P05452 | CLEC3B | P18206 | VCL | P08263 | GSTA1 | A0A0C4DH36 | IGHV3-38 |
|  |  | P06276 | BCHE | P21333 | FLNA | P0C0S8 | H2AC17 | A0A0C4DH38 | IGHV5-51 |
|  |  | P08519 | LPA | P26927 | MST1 | P11021 | HSPA5 | A0A0C4DH67 | IGKV1-8 |
|  |  | P0C0L4 | C4A | P35542 | SAA4 | P11142 | HSPA8 | A0A0C4DH68 | IGKV2-24 |
|  |  | P0C0L5 | C4B_2 | P35579 | MYH9 | P12955 | PEPD | O43866 | CD5L |
|  |  | P10909 | CLU | P43251 | BTD | P13796 | LCP1 | P01591 | JCHAIN |
|  |  | P19823 | ITIH2 | P48740 | MASP1 | P14625 | HSP90B1 | P01593 | IGKV1D-33 |
|  |  | P20742 | PZP | P68363 | TUBA1B | P17174 | GOT1 | P01597 | IGKV1-39 |
|  |  | P20851 | C4BPB | Q08830 | FGL1 | P21549 | AGXT | P01599 | IGKV1-17 |
|  |  | P22891 | PROZ | Q14520 | HABP2 | P26038 | MSN | P01601 | IGKV1D-16 |
|  |  | P27169 | PON1 | Q15942 | ZYX | P29401 | TKT | P01602 | IGKV1-5 |
|  |  | P29622 | SERPINA4 | Q92954 | PRG4 | P30041 | PRDX6 | P01619 | IGKV3-20 |
|  |  | P35858 | IGFALS | Q9BXR6 | CFHR5 | P30101 | PDIA3 | P01700 | IGLV1-47 |
|  |  | P49908 | SELENOP | Q9NZP8 | C1RL | P32119 | PRDX2 | P01701 | IGLV1-51 |
|  |  | P55056 | APOC4 | Q9UK55 | SERPINA10 | P37837 | TALDO1 | P01703 | IGLV1-40 |
|  |  | P80108 | GPLD1 | Q9Y490 | TLN1 | P60709 | ACTB | P01704 | IGLV2-14 |
|  |  | Q03591 | CFHR1 |  |  | P62805 | H4C16 | P01709 | IGLV2-8 |
|  |  | Q04756 | HGFAC |  |  | P63104 | YWHAZ | P01714 | IGLV3-19 |
|  |  | Q13790 | APOF |  |  | P68032 | ACTC1 | P01718 | IGLV3-27 |
|  |  | Q15848 | ADIPOQ |  |  | P68104 | EEF1A1 | P01742 | IGHV1-69 |
|  |  | Q16610 | ECM1 |  |  | P68871 | HBB | P01743 | IGHV1-46 |
|  |  | Q92496 | CFHR4 |  |  | P69905 | HBA2 | P01764 | IGHV3-23 |
|  |  | Q96KN2 | CNDP1 |  |  | P78417 | GSTO1 | P01780 | IGHV3-7 |
|  |  | Q96PD5 | PGLYRP2 |  |  | Q13228 | SELENBP1 | P01782 | IGHV3-9 |
|  |  | Q9UGM5 | FETUB |  |  | Q16851 | UGP2 | P01824 | IGHV4-39 |
|  |  |  |  |  |  |  |  | P01834 | IGKC |
|  |  |  |  |  |  |  |  | P01860 | IGHG3 |
|  |  |  |  |  |  |  |  | P01861 | IGHG4 |
|  |  |  |  |  |  |  |  | P01871 | IGHM |
|  |  |  |  |  |  |  |  | P01876 | IGHA1 |
|  |  |  |  |  |  |  |  | P04211 | IGLV7-43 |
|  |  |  |  |  |  |  |  | P04430 | IGKV1-16 |
|  |  |  |  |  |  |  |  | P06312 | IGKV4-1 |
|  |  |  |  |  |  |  |  | P0DOX2 | NA |
|  |  |  |  |  |  |  |  | P0DOX4 | NA |
|  |  |  |  |  |  |  |  | P0DOX6 | NA |
|  |  |  |  |  |  |  |  | P0DOX7 | NA |
|  |  |  |  |  |  |  |  | P0DOX8 | NA |
|  |  |  |  |  |  |  |  | P0DOY3 | IGLC3 |
|  |  |  |  |  |  |  |  | P15814 | IGLL1 |
|  |  |  |  |  |  |  |  | P23083 | IGHV1-2 |
|  |  |  |  |  |  |  |  | P80748 | IGLV3-21 |
|  |  |  |  |  |  |  |  | A0A075B6H9 | IGLV4-69 |
|  |  |  |  |  |  |  |  | A0A0A0MT36 | IGKV6D-21 |
|  |  |  |  |  |  |  |  | A0A0C4DH25 | IGKV3D-20 |
|  |  |  |  |  |  |  |  | Q68DL7 | C18orf63 |
|  |  |  |  |  |  |  |  | Q9NP80 | PNPLA8 |
| **Day 4** | | | | | | | | | |
|  | | **β pattern** | | **γ pattern** | | **δ pattern** | | **ε pattern** | |
|  |  | **Gene** | **Identifier** | **Gene** | **Gene** | **Identifier** | **Gene** | **Identifier** | **Gene** |
|  |  | O75882 | ATRN | P00738 | HP | O60814 | H2BC12 | A0A075B6H7 | IGKV3-7 |
|  |  | O95445 | APOM | P00751 | CFB | O75874 | IDH1 | A0A075B6I0 | IGLV8-61 |
|  |  | P00488 | F13A1 | P02741 | CRP | P00338 | LDHA | A0A075B6K4 | IGLV3-10 |
|  |  | P00739 | HPR | P02743 | APCS | P00746 | CFD | A0A075B6K5 | IGLV3-9 |
|  |  | P00740 | F9 | P02750 | LRG1 | P00915 | CA1 | A0A075B6P5 | IGKV2-28 |
|  |  | P01024 | C3 | P02775 | PPBP | P01019 | AGT | A0A075B6S5 | IGKV1-27 |
|  |  | P02647 | APOA1 | P07357 | C8A | P02144 | MB | A0A087WSY6 | IGKV3D-15 |
|  |  | P02652 | APOA2 | P0DJI8 | SAA1 | P04040 | CAT | A0A087WSZ0 | IGKV1D-8 |
|  |  | P02654 | APOC1 | P0DJI9 | SAA2 | P04075 | ALDOA | A0A0A0MRZ8 | IGKV3D-11 |
|  |  | P02655 | APOC2 | P18206 | VCL | P04179 | SOD2 | A0A0A0MS15 | IGHV3-49 |
|  |  | P02656 | APOC3 | P21333 | FLNA | P04275 | VWF | A0A0B4J1U3 | IGLV1-36 |
|  |  | P02753 | RBP4 | P35579 | MYH9 | P04406 | GAPDH | A0A0B4J1U7 | IGHV6-1 |
|  |  | P02766 | TTR | P61224 | RAP1B | P05062 | ALDOB | A0A0B4J1V0 | IGHV3-15 |
|  |  | P02790 | HPX | P68363 | TUBA1B | P05109 | S100A8 | A0A0B4J1V2 | IGHV2-26 |
|  |  | P03951 | F11 | Q15942 | ZYX | P06702 | S100A9 | A0A0B4J1X5 | IGHV3-74 |
|  |  | P03952 | KLKB1 | Q9BXR6 | CFHR5 | P06733 | ENO1 | A0A0B4J1Y8 | IGLV9-49 |
|  |  | P04004 | VTN | Q9UK55 | SERPINA10 | P07237 | P4HB | A0A0B4J1Y9 | IGHV3-72 |
|  |  | P04070 | PROC | Q9Y490 | TLN1 | P07900 | HSP90AA1 | A0A0B4J2D9 | IGKV1D-13 |
|  |  | P04180 | LCAT |  |  | P0C0S8 | H2AC17 | A0A0C4DH24 | IGKV6-21 |
|  |  | P04196 | HRG |  |  | P10643 | C7 | A0A0C4DH31 | IGHV1-18 |
|  |  | P04278 | SHBG |  |  | P11021 | HSPA5 | A0A0C4DH38 | IGHV5-51 |
|  |  | P05154 | SERPINA5 |  |  | P11142 | HSPA8 | A0A0C4DH67 | IGKV1-8 |
|  |  | P05156 | CFI |  |  | P12955 | PEPD | A0A0C4DH68 | IGKV2-24 |
|  |  | P05160 | F13B |  |  | P13796 | LCP1 | O43866 | CD5L |
|  |  | P05452 | CLEC3B |  |  | P14625 | HSP90B1 | P01591 | JCHAIN |
|  |  | P05546 | SERPIND1 |  |  | P17174 | GOT1 | P01593 | IGKV1D-33 |
|  |  | P07358 | C8B |  |  | P21549 | AGXT | P01597 | IGKV1-39 |
|  |  | P0C0L4 | C4A |  |  | P24158 | PRTN3 | P01599 | IGKV1-17 |
|  |  | P0C0L5 | C4B_2 |  |  | P26038 | MSN | P01601 | IGKV1D-16 |
|  |  | P10909 | CLU |  |  | P29401 | TKT | P01602 | IGKV1-5 |
|  |  | P12259 | F5 |  |  | P30041 | PRDX6 | P01619 | IGKV3-20 |
|  |  | P20742 | PZP |  |  | P30101 | PDIA3 | P01700 | IGLV1-47 |
|  |  | P20851 | C4BPB |  |  | P32119 | PRDX2 | P01701 | IGLV1-51 |
|  |  | P22352 | GPX3 |  |  | P36222 | CHI3L1 | P01703 | IGLV1-40 |
|  |  | P26927 | MST1 |  |  | P37837 | TALDO1 | P01704 | IGLV2-14 |
|  |  | P27169 | PON1 |  |  | P51884 | LUM | P01709 | IGLV2-8 |
|  |  | P29622 | SERPINA4 |  |  | P60709 | ACTB | P01714 | IGLV3-19 |
|  |  | P35858 | IGFALS |  |  | P61769 | B2M | P01718 | IGLV3-27 |
|  |  | P48740 | MASP1 |  |  | P62805 | H4C16 | P01742 | IGHV1-69 |
|  |  | P49908 | SELENOP |  |  | P63104 | YWHAZ | P01743 | IGHV1-46 |
|  |  | P55056 | APOC4 |  |  | P68032 | ACTC1 | P01764 | IGHV3-23 |
|  |  | P80108 | GPLD1 |  |  | P68871 | HBB | P01780 | IGHV3-7 |
|  |  | Q04756 | HGFAC |  |  | P69905 | HBA2 | P01782 | IGHV3-9 |
|  |  | Q13790 | APOF |  |  | P78417 | GSTO1 | P01824 | IGHV4-39 |
|  |  | Q14520 | HABP2 |  |  | Q16851 | UGP2 | P01833 | PIGR |
|  |  | Q15848 | ADIPOQ |  |  | O43286 | B4GALT5 | P01834 | IGKC |
|  |  | Q92954 | PRG4 |  |  | P07339 | CTSD | P01860 | IGHG3 |
|  |  | Q96IY4 | CPB2 |  |  | Q9Y279 | VSIG4 | P01861 | IGHG4 |
|  |  | Q96PD5 | PGLYRP2 |  |  |  |  | P01871 | IGHM |
|  |  | Q9UGM5 | FETUB |  |  |  |  | P01876 | IGHA1 |
|  |  | P49913 | CAMP |  |  |  |  | P04211 | IGLV7-43 |
|  |  |  |  |  |  |  |  | P04430 | IGKV1-16 |
|  |  |  |  |  |  |  |  | P05362 | ICAM1 |
|  |  |  |  |  |  |  |  | P06312 | IGKV4-1 |
|  |  |  |  |  |  |  |  | P0DOX2 | NA |
|  |  |  |  |  |  |  |  | P0DOX4 | NA |
|  |  |  |  |  |  |  |  | P0DOX6 | NA |
|  |  |  |  |  |  |  |  | P0DOX7 | NA |
|  |  |  |  |  |  |  |  | P0DOX8 | NA |
|  |  |  |  |  |  |  |  | P0DOY3 | IGLC3 |
|  |  |  |  |  |  |  |  | P15814 | IGLL1 |
|  |  |  |  |  |  |  |  | P19320 | VCAM1 |
|  |  |  |  |  |  |  |  | P23083 | IGHV1-2 |
|  |  |  |  |  |  |  |  | P35527 | KRT9 |
|  |  |  |  |  |  |  |  | P35908 | KRT2 |
|  |  |  |  |  |  |  |  | P80748 | IGLV3-21 |
|  |  |  |  |  |  |  |  | Q9Y6R7 | FCGBP |
|  |  |  |  |  |  |  |  | A0A0A0MT36 | IGKV6D-21 |
|  |  |  |  |  |  |  |  | A0A0C4DH25 | IGKV3D-20 |
|  |  |  |  |  |  |  |  | Q9Y5Y7 | LYVE1 |

**Supplementary Figures**

**Supplementary Figure 1: Quality control for proteomics data normalization. a** Principal component analysis (PCA) plots of mass spectrometry data before and after batch normalization. Each data point corresponds to a sample, colors representing the respective batches. **b** Boxplot representation of protein intensities before and after normalization. Boxes indicate the 25% - 75% interquartile range (IQR) with the median displayed as horizontal line. Whiskers extend to 1.5 x IQR. Outliers displayed as individual data points. Each box corresponds to a sample, colors representing the respective batches.

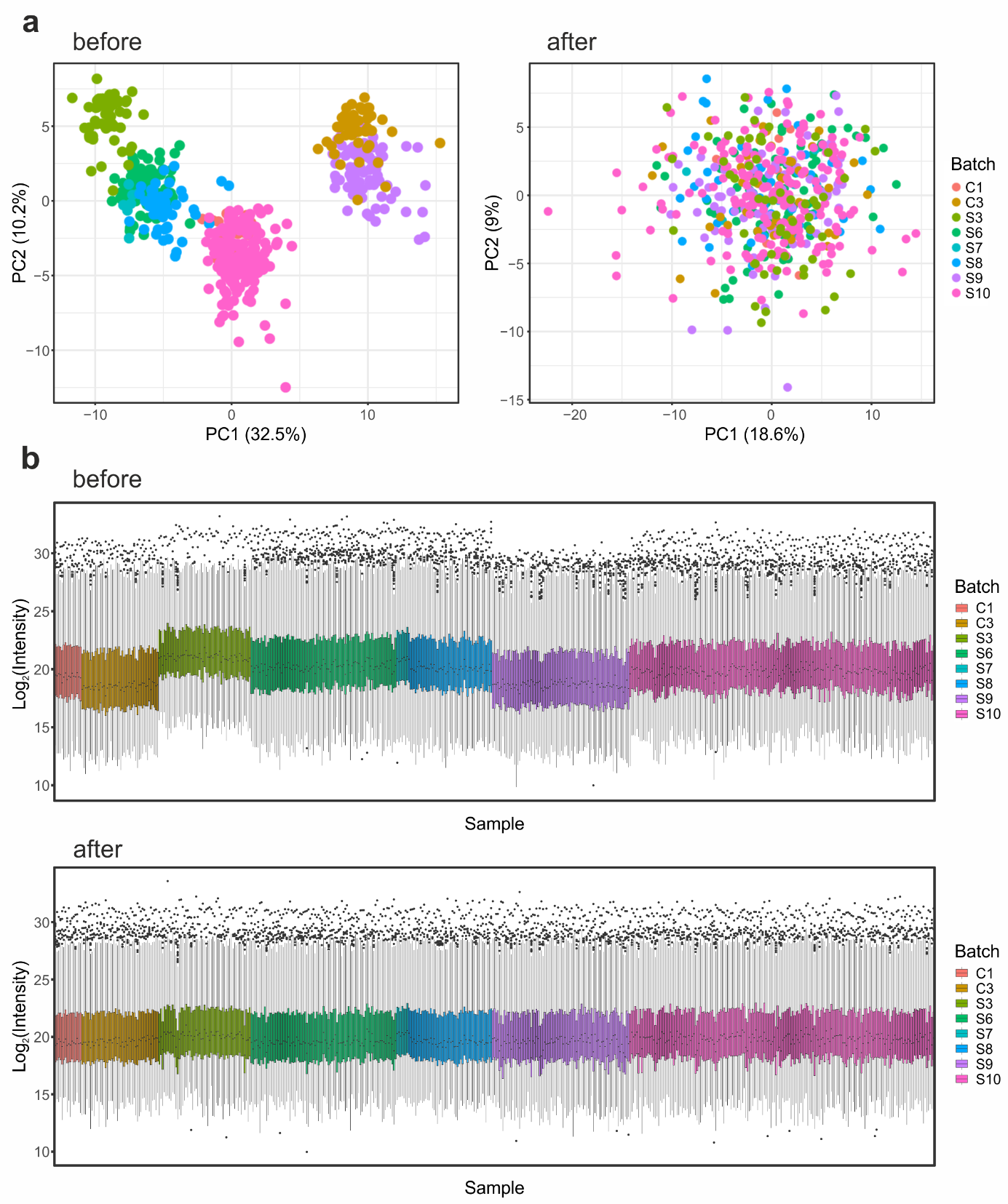

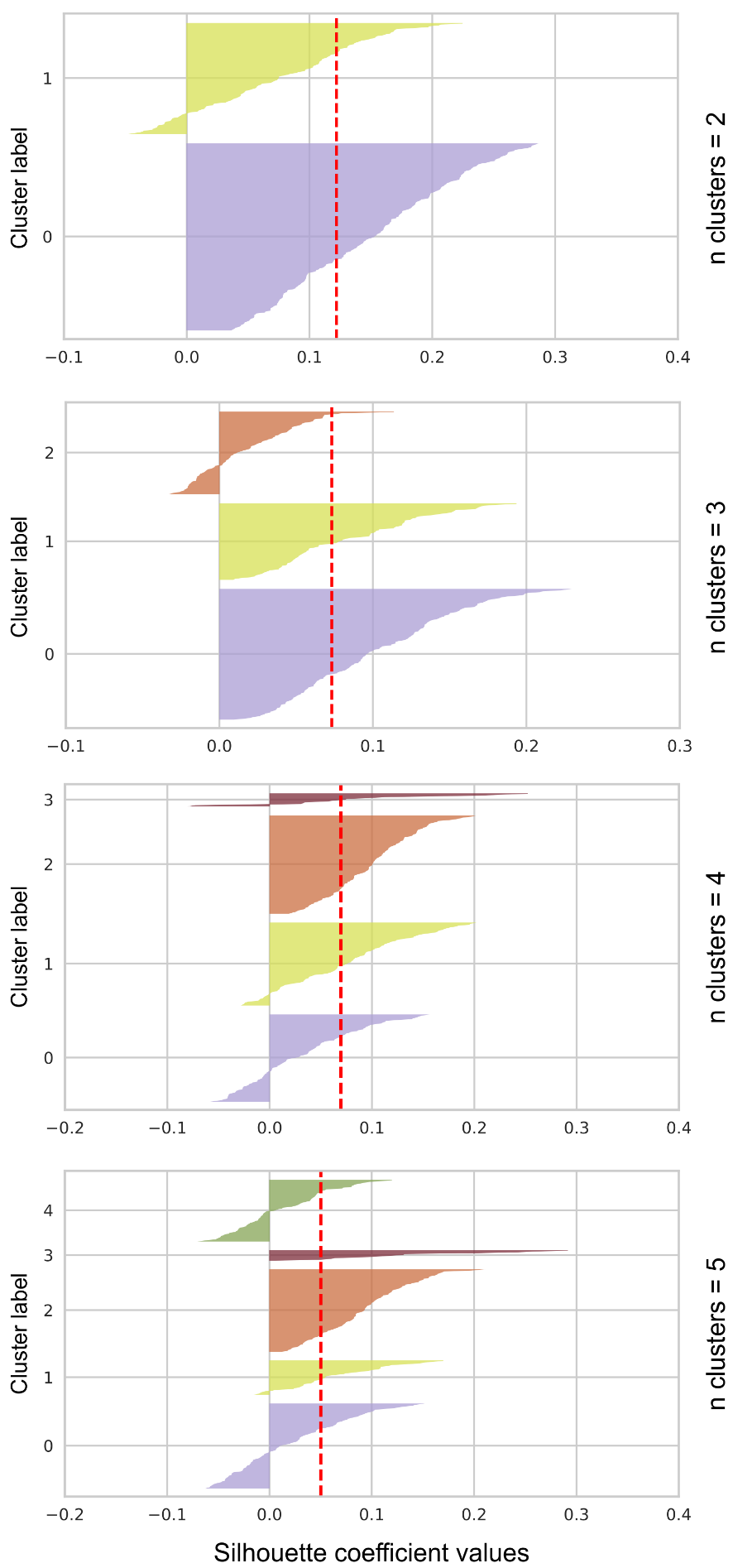

**Supplementary Figure 2: Silhouette curves that were used to determine of the optimal number of clusters.** Red dotted line representing the mean silhouette coefficient. While two cluster yielded the highest average silhouette score, it lacked sufficient resolution between subgroups. Three cluster showed a noticeable drop in silhouette values. The 5-cluster solution provided slightly improved separation for some clusters, but the number of points in the cluster decreased. Four cluster offered the best trade-off between cohesion and separation and was also the most biologically meaningful, supporting its selection for downstream analyses. For the chosen 4 clusters the labeling was changed to represent the gradual severity of sepsis.

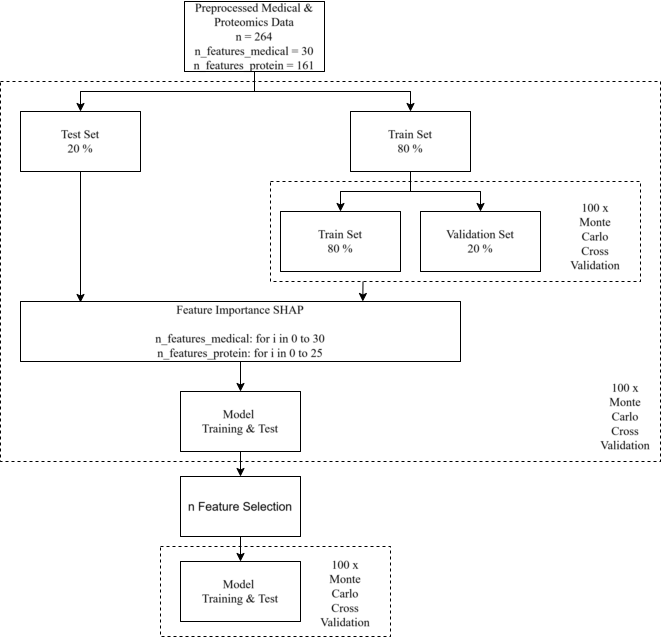

**Supplementary Figure 3: Framework for machine learning and feature selection.** Schematic representation of the applied workflow for training of rando forest classifiers and selection of the most relevant features.

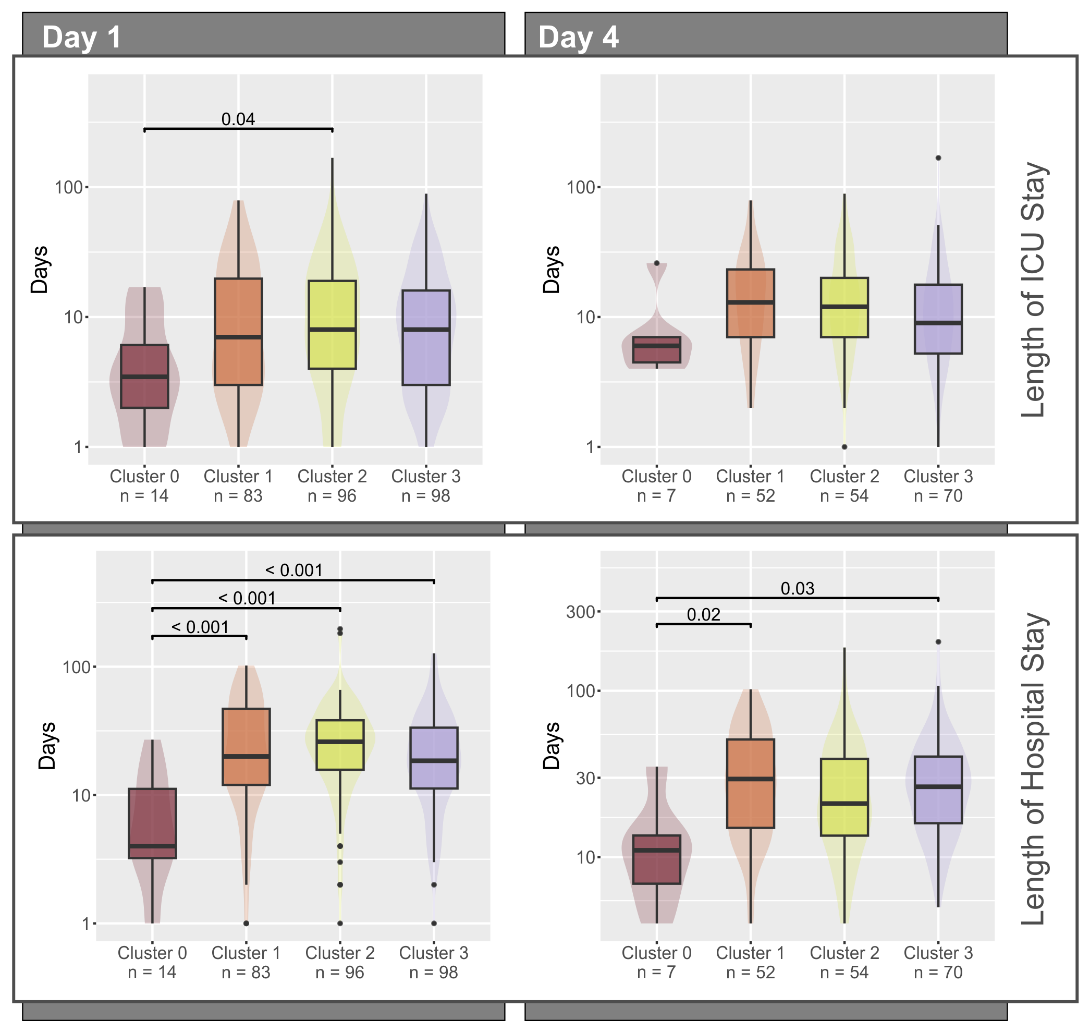

**Supplementary Figure 4: Time of ICU stay and hospital stay.** Times of ICU stay and hospital stay as boxplot representation separated for days 1 and 4 and the four identified molecular subtypes. Boxes represent 25th and 75th percentiles, whiskers extend to the most extreme data points, median shown as a horizontal line, outliers shown as individual data points, p-values calculated by Dunn’s post-hoc test.

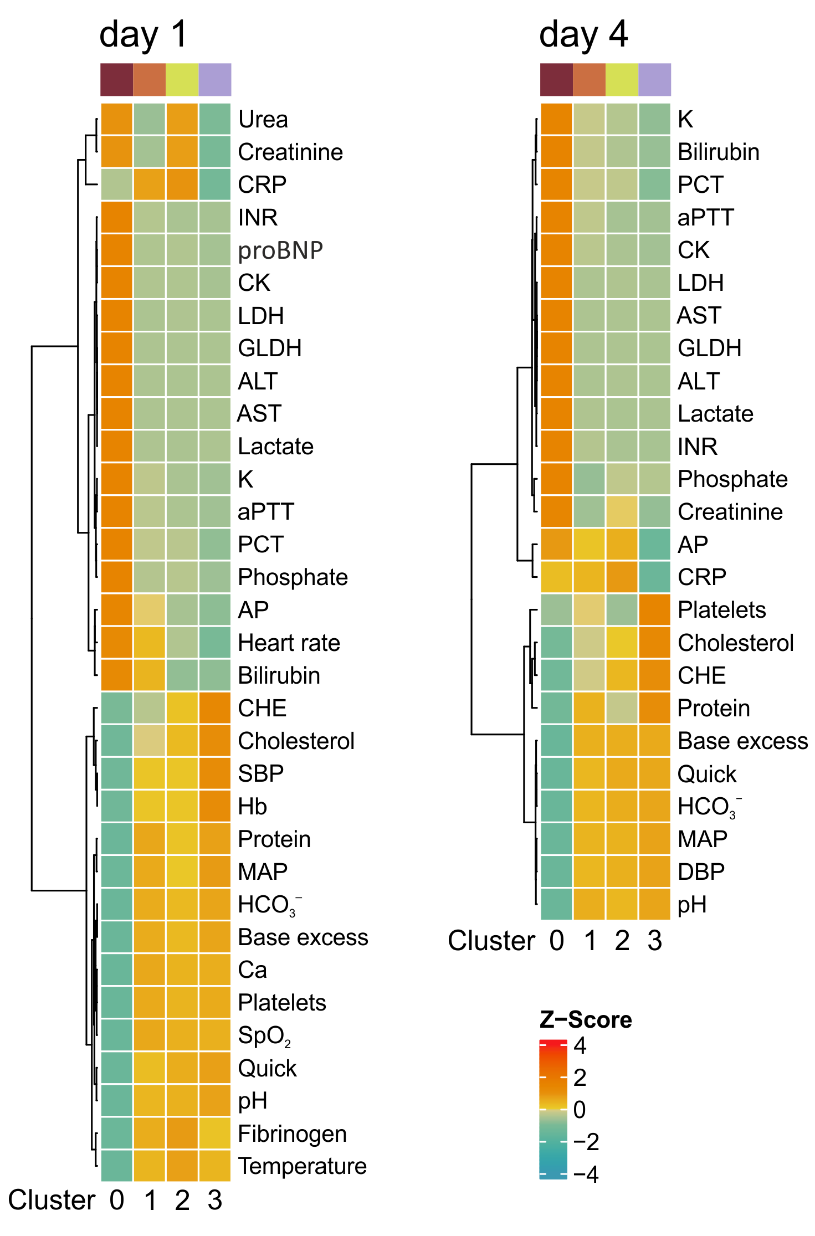

**Supplementary Figure 5: Clinical characteristics of the sepsis plasma proteome subtypes.** Heatmaps illustrating clinical routine data at day 1 and day 4. All parameters displayed which were significant in any pairwise comparison between the subtypes (Bonferroni corrected Kruskal-Wallis followed by Dunn's test). Data was z-transformed, and clustered using Euclidean distance with Ward's linkage method.

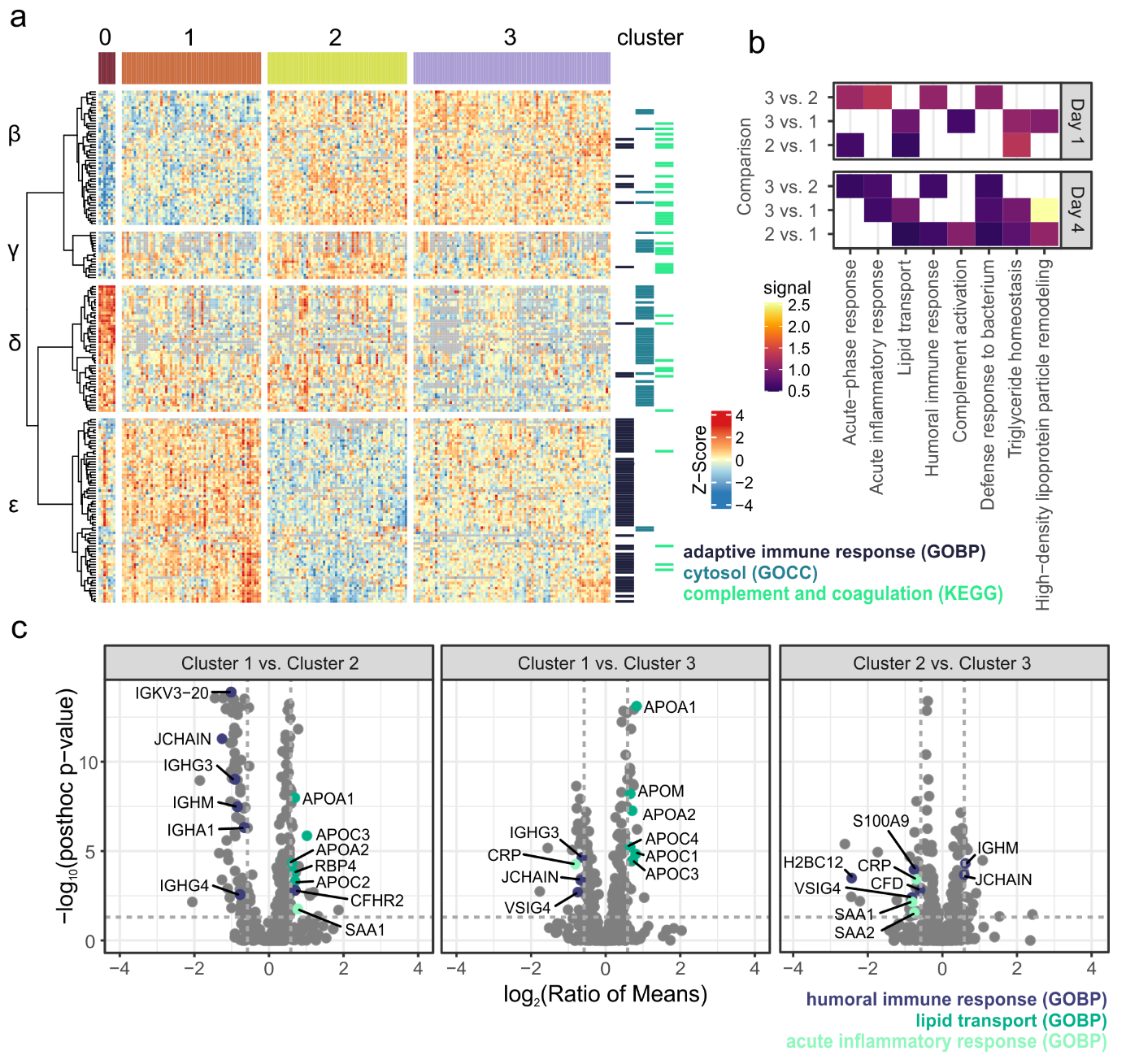

**Supplementary Figure 6: Proteome characteristics of the sepsis plasma proteome subtypes at day 4. a** Heatmap showing all significantly differentially abundant proteins at day 4 (ANOVA pFDR value ≤ 0.05, post-hoc test p value ≤ 0.05, ratio of means ≥ 1.5 or ≤ 0.67). Protein intensities were z-transformed and clustered using Pearson clustering with Ward's linkage method. The first four branches in the dendrogram were divided and labeled with Greek letters to discriminate abundance patterns. Protein annotation with selected gene ontology or KEGG categories is indicated on the right. **b** Functional enrichment of all significantly proteins significantly differentially abundant between clusters 1, 2 and 3. Eight selected categories shown for day 1 and day 4 and the three pairwise comparisons. Enrichment analysis was done with string-db.org (v.12) using GO biological processes (GOBP). **c** Volcano plots illustrating the pairwise comparisons between clusters 1, 2 and 3 for day 1. Proteins annotated with significantly enriched GOBP terms were highlighted and labeled with gene names. Dashed lines indicate the applied significance threshold.

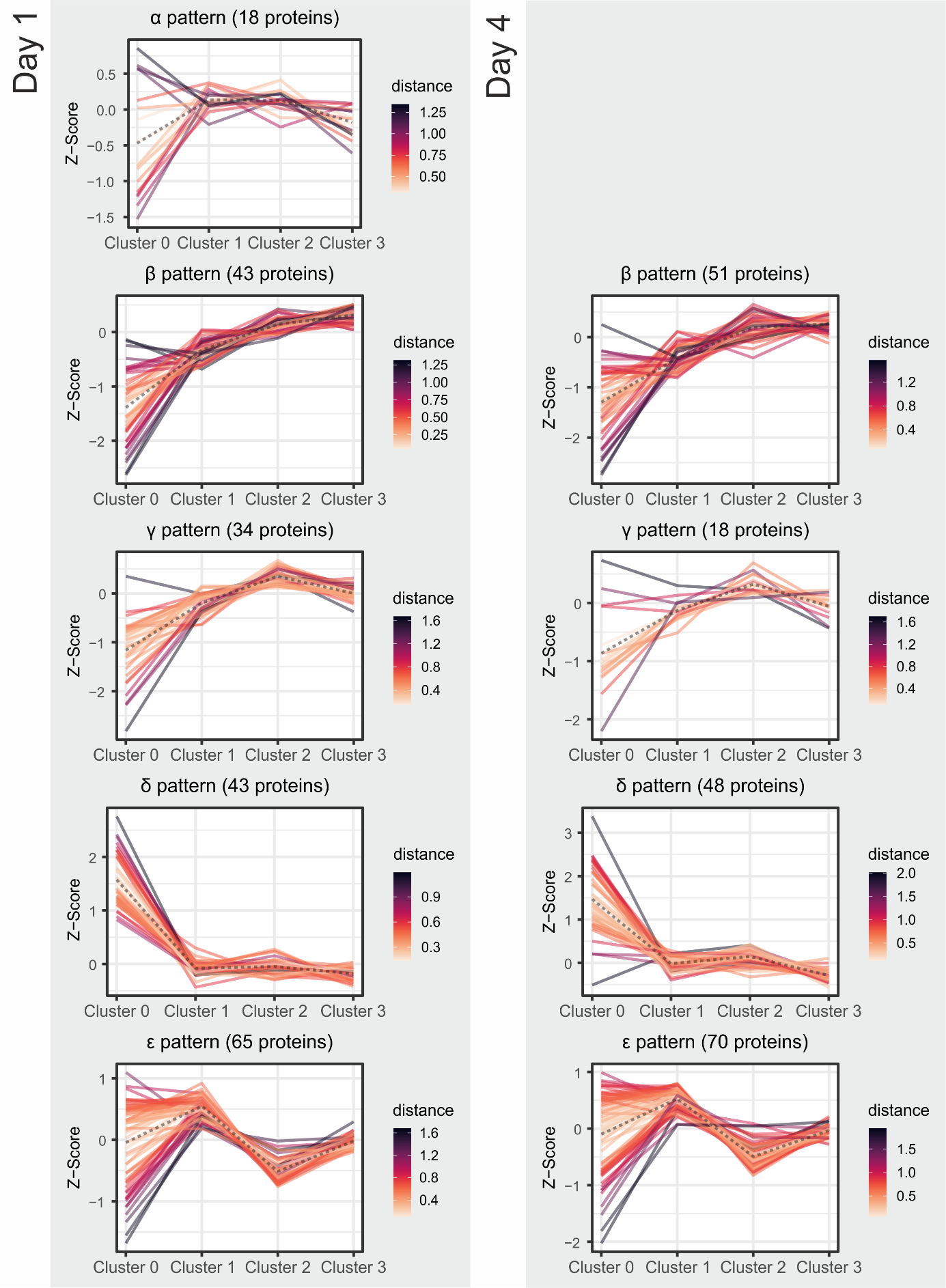

**Supplementary Figure 7: Identified protein abundance patterns for day 1 and day 4.** Line plots showing z-scored mean protein abundances for each protein and the four plasma proteome subtypes. The centre of each pattern is displayed as dashed line and the Euclidean distance to the centre is colour-coded for each protein. The α pattern was not observed on day 4.

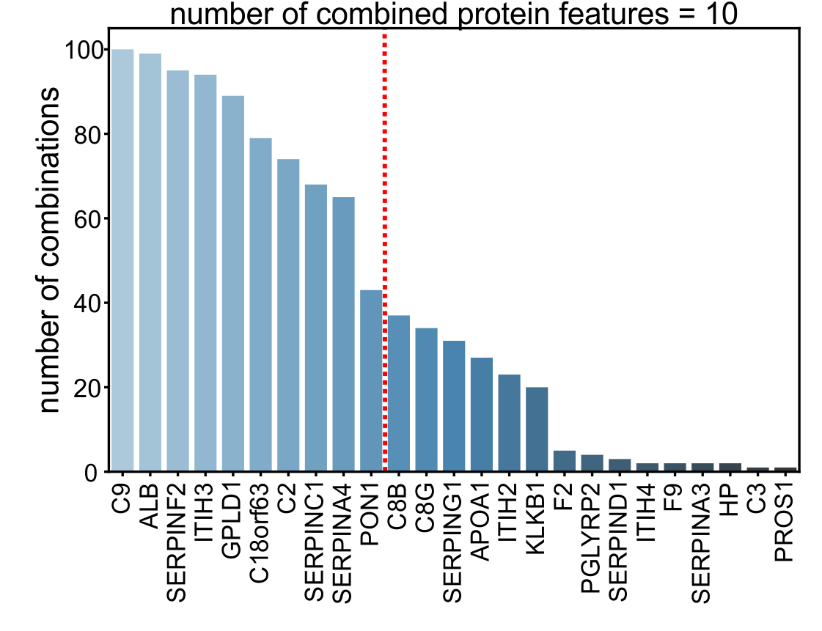

**Supplementary Figure 8: Feature Selection for the machine learning classifier.** Bar chart showing the frequency at which a feature was included in a combination of ten protein and no medical features during MCCV. Red line indicating the threshold of 10 proteins which were selected for training of the final model.

**Supplementary Figure 9: SHAP summary plots for the machine learning classifier.** Scatter plots showing the impact of each feature on the model's prediction over all MCCV iterations. Colors indicate feature values (blue = low, pink = high). Data shown separately for prediction of clusters 1, 2 and 3.

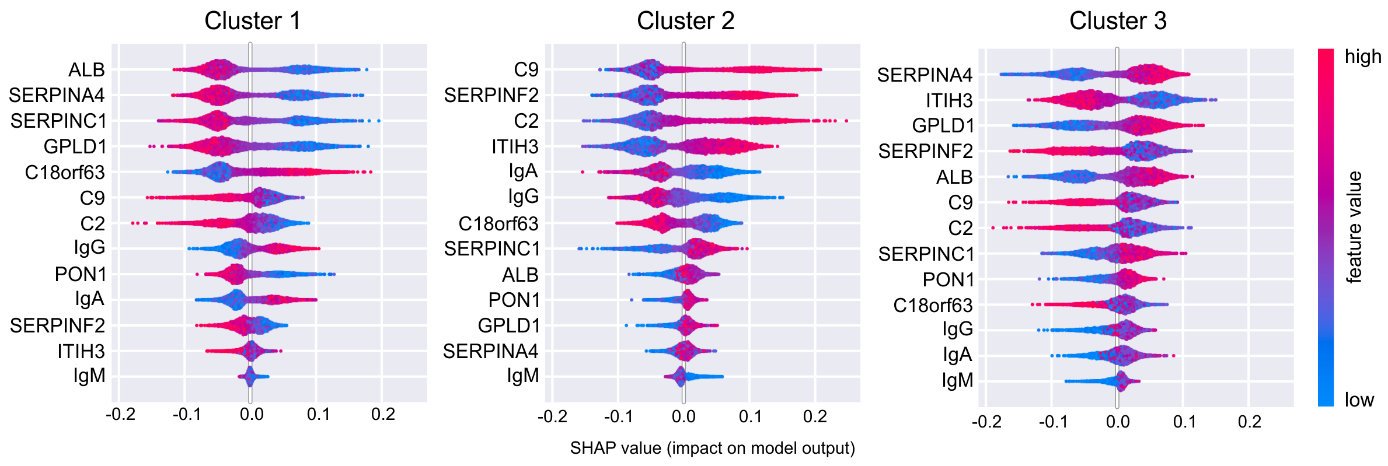
